# Identifying Sex-Specific Sub-phenotypes of Alzheimer’s Disease Progression Using Longitudinal Electronic Health Records

**DOI:** 10.1101/2024.07.07.24310055

**Authors:** Weimin Meng, Qiang Yang, Jie Xu, Yu Huang, Cankun Wang, Qianqian Song, Lixin Song, Jiang Bian, Qin Ma, Anjun Ma, Rui Yin

## Abstract

Alzheimer’s Disease (AD) is a complex neurodegenerative disorder strongly influenced by sex differences, with women comprising nearly two-thirds of cases. However, sex-specific progression patterns remain underexplored due to unclear clinical and molecular mechanisms. To address this gap, we developed a temporal autoencoder framework to identify sex-specific AD sub-phenotypes using longitudinal electronic health record (EHR) data from the OneFlorida+ Clinical Research Consortium. Sequential EHRs were encoded into latent representations and clustered to derive disease states, which were assembled into progression pathways. This approach uncovered five primary sex-stratified sub-phenotypes with distinct trajectories and phenotypic characteristics. Survival and cumulative prevalence analyses further revealed heterogeneous temporal dynamics of AD onset and comorbidity accumulation between female- and male-dominant groups. By integrating deep learning with large-scale real-world data, our framework advances understanding of sex-based heterogeneity in AD progression and provides a scalable tool for early risk stratification, personalized intervention, and improved clinical trial design.

## Introduction

Alzheimer’s disease (AD), recognized as the predominant form of dementia, is a multifaceted and progressive neurodegenerative disorder that currently affects an estimated 6.9 million Americans as of 2024^1^, with a potential rise to 13.85 million by 2060^2^. AD accounts for 60-80% of dementia cases as well as the 7^th^ leading cause of death in the United States, which imposes substantial burdens on individuals affected, their families, healthcare systems, and the whole society. The progression of AD is hypothesized to include three main phases: preclinical AD, clinically significant mild cognitive impairment (MCI), and AD, forming a continuum of increasing cognitive decline^3^. Early pathological changes, such as beta-amyloid plaques and tau tangles, begin in the preclinical stage^4^, leading to MCI with cognitive decline and eventually severe cognitive deficits, memory loss, and functional impairment in AD^5^. However, the progression of AD is characterized by a complex array of longitudinally linked clinical features and outcomes, with a broad spectrum of manifestations beyond the three main phases^6–8^. There is considerable interindividual variability in AD progression^9^, with some experiencing rapid loss of cognition while others progress more slowly. Compelling evidence indicates that heterogeneity exists in AD progression through different intermediate stages with varied clinical presentations^10–13^. The considerable variability^7^ in AD progression^9^ and clinical phenotypes^10–12^ presents a challenge in understanding how these factors influence the disease trajectory.

Basic and clinical research have indicated that sex differences are one of the most critical factors contributing to its complexity^14–17^. Approximately two-thirds of all existing AD cases are in females^1^, and recent studies suggest significant sex differences in clinical severity^18–20^, neuropathological characteristics^21,22^, and genetic factors^15,23^ in AD. It is also reported that for women aged 65, the lifetime risk of developing AD is 21.2%, about twice the risk seen in men^1,24^. Additionally, female sex is a major risk factor for late-onset AD^25^. One of the reasons for the higher prevalence of AD in women might be their longer average life expectancy. Regarding AD progression, accelerated cognitive decline in females has partially been attributed to the effects of the APOE ε4 allele^25–28^. APOE ε4 significantly increases brain Aβ deposition and atrophy and dramatically reduces brain connectivity in the default mode network of females^29,30^. Several biomarkers have been revealed (e.g., Cerebrospinal Fluid, neuroimage), providing clearer evidence of a negative correlation between the APOE ε4 genotype and AD in female patients^31^. In terms of clinical phenotypes, some studies found that women are more likely to develop multiple comorbidities, which may interact and collectively increase their risk of developing AD^32^. However, despite these pathological changes and comorbidity interactions, other existing evidence suggested that women have greater cognitive resilience^33^. It is reported that women might be better able to preserve their brain structural properties after exposure to pathological tau^34^. The mounting proof of sex differences in AD highlights the importance of understanding the underlying mechanisms and AD progression in females and males. Despite substantial research on sex differences in AD, the heterogeneity of sex-specific progression patterns within distinct AD sub-phenotypes, defined as subtypes characterized by phenotypic features^12,35–38^, remains under-investigated.

The longitudinal electronic health records (EHRs)^39,40^ consist of a wide variety of critical health events of patients through routine care, including diagnoses, comorbidities, medications, laboratory measurements, and other relevant clinical information, which can offer long-term insights into AD development^41–43^. These longitudinal EHRs have been effectively utilized to predict patient outcomes and identify disease sub-phenotypes, including AD. For example, Lee et al.^44^ developed deep predictive clustering on temporal phenotyping to identify distinct AD subgroups with varying disease trajectories, including those with faster cognitive decline, higher comorbidity burdens (e.g., cardiovascular diseases, diabetes), and differential responses to treatment interventions. Xu et al.^42^ proposed an outcome-oriented model using Long Short-Term Memory (LSTM)^45^ to identify progression pathways for AD onset, deriving several AD progression subtypes related to clinical phenotypes such as cardiovascular diseases. In terms of sex difference study of AD, Tang et al.^46^ performed comprehensive phenotyping and network analyses, gaining insight into clinical characteristics and sex-specific clinical associations in AD. Furthermore, Tang et al.^47^ demonstrated how EHRs and knowledge networks can be leveraged for AD onset prediction from different index times and uncover sex-specific biological insights. However, the role of sex in shaping the complexity and heterogeneity of AD progression remains largely unexplored. Existing approaches often overlook how sex influences the long-term pathological trajectories throughout the course of AD^38^. Characterizing sex-specific progression patterns or sub-phenotypes in AD and identifying their contributing factors is essential for advancing personalized strategies to optimize AD prevention and care. A deeper understanding of sex differences can enable clinicians to customize therapeutic strategies more effectively, maximizing treatment benefits while minimizing adverse effects for individuals living with AD.

In this study, we developed an autoencoder architecture to identify sex-specific AD progression sub-phenotypes using large-scale longitudinal EHRs from the OneFlorida+ clinical research network. Unlike traditional approaches that analyze associations between clinical characteristics and patient outcomes, our autoencoder architecture can capture dynamic disease progression while accounting for sex stratification. Specifically, we first identified and collected longitudinal EHRs of AD cohorts from the OneFlorida+, constructing patient-specific temporal matrices to represent sequential clinical records. A novel autoencoder was developed involving a three-layer encoder and a three-layer decoder to generate outcome-oriented latent representations that reflect disease trajectory. Using hierarchical agglomerative clustering methods, we grouped latent embeddings into distinct clusters representing different disease states. We further analyzed and compared the distributions of phenotypic features across clusters and identified sex-specific AD sub-phenotypes based on major state transitions within patients’ progression pathways. These sex-specific sub-phenotypes were interpreted through phenotype-based statistical analysis and progression modeling. This framework provides new insights into the heterogeneity of AD progression in terms of sex differences, potentially aiding in personalized AD treatment and care. The overall workflow of this framework is presented in Fig. 1.

**Fig. 1:**
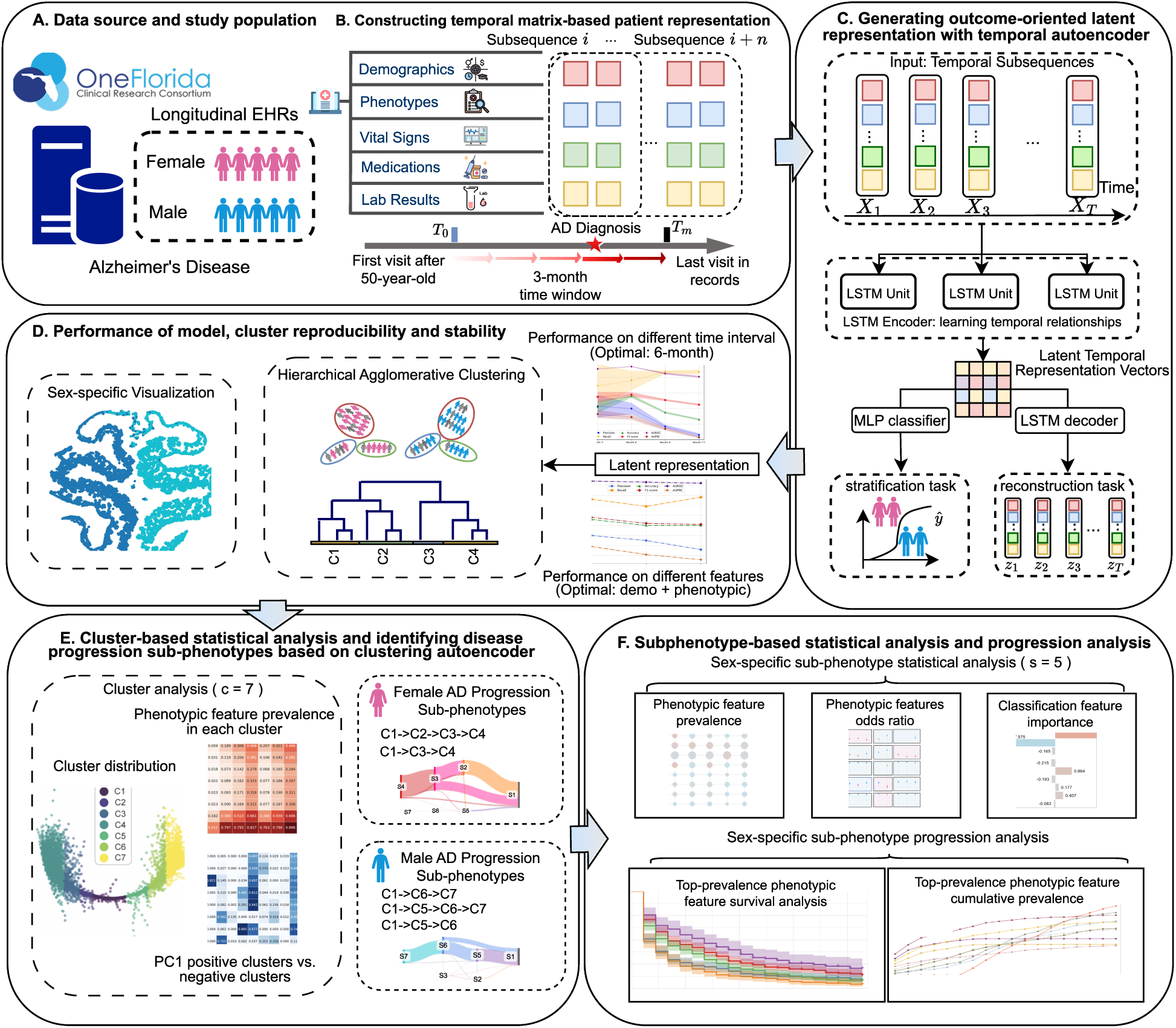
The overview of the study. **A**. Data source and cohort identification. **B**. The construction of longitudinal EHRs for individuals with AD. **C**. Sex-specific AD latent representation generation with our proposed autoencoder framework. **D**. Model optimization and evaluation. **E**. Sex-specific AD progression sub-phenotypes identification and characterization. **F**. Sex-specific AD progression sub-phenotypes analysis and interpretation.

## Results

### Cohort summary and characteristics

As shown in Table 1, we identified 1,665 individuals with AD from the OneFlorida+ Clinical Research Network, consisting of 961 females (mean age at first encounter: 76.7 ± 9.2 years) and 704 males (mean age at first encounter: 76.1 ± 8.8 years). According to the table, we found that females made up a slightly larger proportion than males (57.7% versus 42.3%) and were diagnosed at an older average age (77.1 years versus 76.1 years). They had a longer mean follow-up duration than males (8.21 years vs. 7.95 years). Meanwhile, the average years before diagnosis was nearly the same between females and males (5.59 ± 1.73 years vs. 5.6 ± 1.7 years), whereas females had a longer post-diagnosis duration than males (2.62 ± 1.59 years vs. 2.36 ± 1.48 years). This is probably because female individuals with AD have a longer life expectancy, which suggests a slightly higher risk of developing AD. Regarding mortality rates, males show a higher mortality rate (14.20%) compared to females (11.34%). We also observed racial and ethnic disparities, including a higher proportion of Hispanic individuals among females (25.81%) than males (18.52%), and a greater percentage of Black or African American individuals in females (16.3%) compared to males (14.5%). Conversely, a larger proportion of white individuals was found among males (79.12%) relative to females (77.11%). Males have a greater mortality rate (14.20%) than females (11.34%). More details of other demographic and phenotypic characteristics of the cohort can be found in Table 1 and Supplementary Table 2.

**Table 1.** Descriptive statistics on the characteristics of the study cohort.

| Characteristics | Total individuals with AD (N = 1,665) | Females with AD (N = 961) | Males with AD (N = 704) |
| --- | --- | --- | --- |
| **Demographics** |  |  |  |
| Age at AD diagnosis, mean (std) | 76.7 (9.2) | 77.1 (9.5) | 76.1 (8.8) |
| Female, N (%) | 961 (57.7%) | 961 (100.0%) | 0 (0.0%) |
| Male, N (%) | 704 (42.3%) | 0 (0.0%) | 704 (100.0%) |
| **Disease development, mean years (std)** |  |  |  |
| Follow-up durations | 8.1 (1.70) | 8.21 (1.68) | 7.95 (1.72) |
| Years before diagnosis | 5.6 (1.72) | 5.59 (1.73) | 5.6 (1.7) |
| Years after diagnosis | 2.5 (1.55) | 2.62 (1.59) | 2.36 (1.48) |
| Mortality rate, N (%) | 209 (12.55%) | 109 (11.34%) | 100 (14.20%) |
| **Hispanic, N (%)** |  |  |  |
| Hispanic | 426 (25.6%) | 248 (25.8%) | 178 (18.5%) |
| Not Hispanic | 1,230 (73.9%) | 707 (73.6%) | 523 (54.4%) |
| No Hispanic information | 9 (0.5%) | 6 (0.6%) | 3 (0.3%) |
| **Race, N (%)** |  |  |  |
| American Indian or Alaska Native | 0 (0.0%) | 0 (0.0%) | 0 (0.0%) |
| Asian | 10 (0.6%) | 5 (0.5%) | 5 (0.7%) |
| Black or African American | 259 (15.6%) | 157 (16.3%) | 102 (14.5%) |
| White | 1,298 (77.96%) | 741 (77.11%) | 557 (79.12%) |
| Multiple race | 15 (0.90%) | 6 (0.62%) | 9 (1.28%) |
| Unknown | 83 (5.0%) | 52 (5.4%) | 31 (4.40%) |

### Model performance of sex-stratified patients with AD classification

For the evaluation of our proposed framework in identifying sex-stratified patients with AD, we compared distinct autoencoder-based deep learning models, including LSTM autoencoder (LSTMAuto), Transformer^48^ autoencoder (TransformerAuto), gated recurrent units (GRU)^49^ autoencoder (GRUAuto) and multilayer perceptron (MLP)^50^ autoencoder (MLPAuto) against conventional machine learning benchmarks. Model performance was assessed using several standard metrics. As illustrated in Fig. 2A, the autoencoder-based models, specifically, LSTMAuto, TransformerAuto, and GRUAuto demonstrated high and consistent performance, exceeding 90% across all evaluation metrics, which is superior than benchmark machine learning models (best-performing: Decision Tree [DT], AUROC: 0.954; 95% CI: 0.953, 0.954, and MLPAuto, AUROC: 0.635; 95% CI: 0.634–0.635). Notably, GRUAuto achieved the highest AUROC of 0.995 (95% CI: 0.995–0.995), followed closely by LSTMAuto (AUROC: 0.994; 95% CI: 0.994–0.994) and TransformerAuto (AUROC: 0.994; 95% CI: 0.994–0.994). While the performance differences among the RNN-based autoencoders LSTMAuto and GRUAuto, and the Transformer-based autoencoder TransformerAuto, were minimal and statistically negligible, we ultimately selected LSTMAuto for downstream tasks to maintain consistency with prior studies^42^ and simplify model interpretation. We further examined the impact of temporal granularity by evaluating different time intervals (i.e., 3, 6, 9, and 12 months) used to segment the longitudinal EHR data of patients with AD into temporal subsequences. The results are shown in Fig. 2B, where we can observe that the 6-month interval yielded the best overall performance in terms of accuracy (0.956), F1-score (0.958), and AUROC (0.994).

**Fig. 2:**
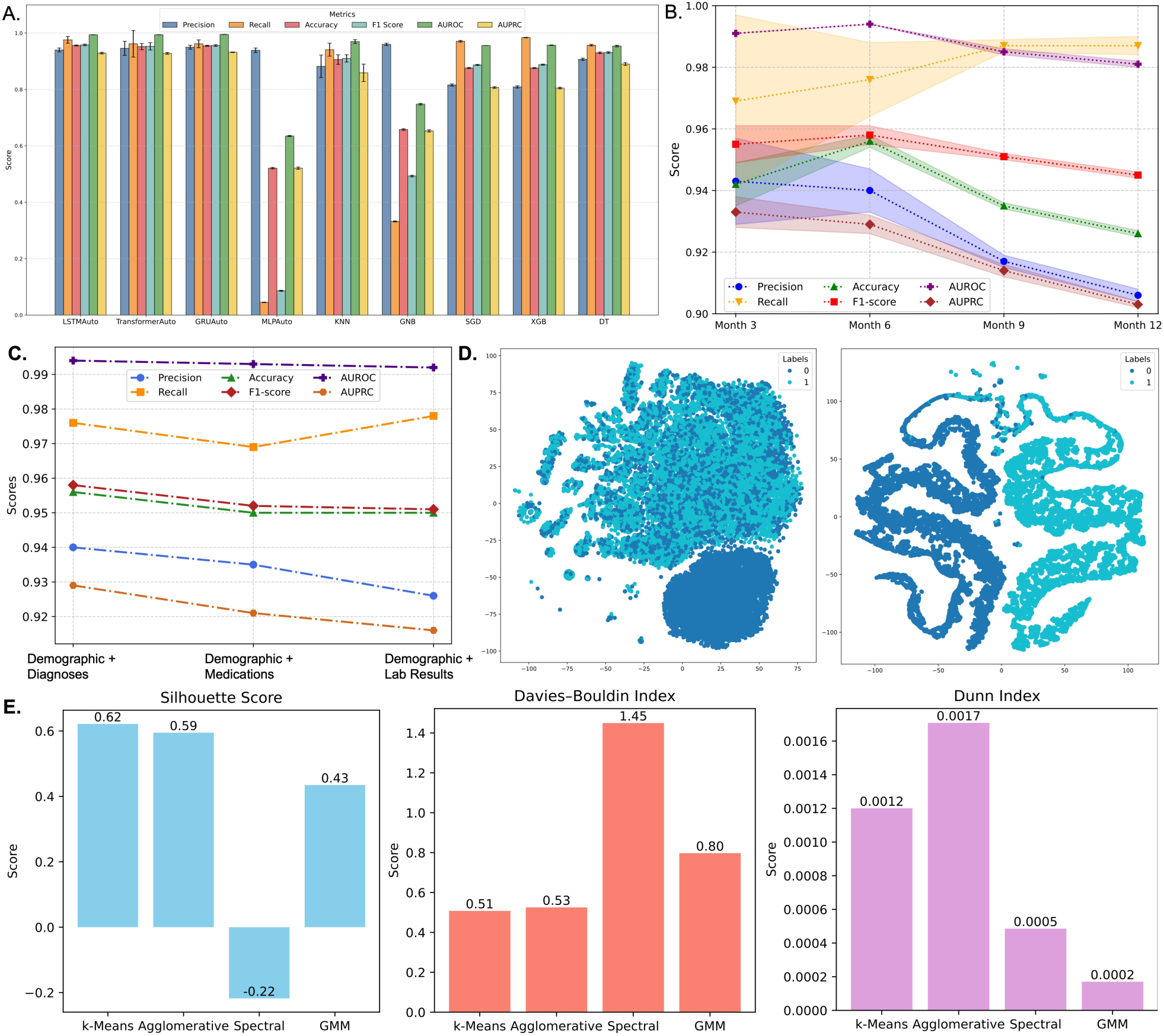
Comparative performance of sex-stratified patients with AD classification and clustering. **A**. The performance of sex-stratified AD classification with our proposed framework and other baselines. **B**. The performance of the LSTM Autoencoder across different time points (Month 3, 6, 9, and 12) under different metrics. The plot shows the mean ± standard deviation (shaded areas), where larger shaded areas indicate higher variability (standard deviation) across measurements. **C**. The impact of different EHR categories on the performance of the LSTM Autoencoder under different metrics. The model was evaluated using three feature sets: Demographic + Diagnoses, Demographic + Medications, and Demographic + Lab Results. **D**. The comparison of t-SNE visualizations of original (left) and learned (right) embeddings for patients with AD. Each point represents a patient with AD, colored by sex label (“0” = male, “1” = female). **E**. The evaluation of different clustering algorithms (k-Means, Agglomerative, Spectral, and GMM) applied to sex-stratified subsequences of patients with AD. Clustering performance is assessed using three internal validation metrics: Silhouette Score (higher is better), Davies–Bouldin Index (lower is better), and Dunn Index (higher is better), reflecting cluster compactness and separation.

In contrast, the 3-month interval produced the highest precision (0.943) and AUPRC (0.991), while the 12-month interval achieved the best recall (0.987). As a result, we will use the LSTMAuto with a 6-month time interval for subsequent analysis due to its optimal performance over others. Furthermore, we conducted a comparative analysis of different combinations of longitudinal EHR data for identifying sex-stratified patients with AD. To evaluate the predictive power of different EHR categories, we trained models for prediction using combinations of demographic information and longitudinal EHR data, including diagnoses, medications, and laboratory results. Fig. 2C shows that the combination of demographics and diagnoses information achieved the highest AUROC (0.994), slightly surpassing demographics with medications (AUROC = 0.993) and demographics with lab results (AUROC = 0.992).

To validate the effectiveness of the LSTMAuto model in distinguishing sex-stratified patients with AD, we employed t-distributed stochastic neighbor embedding (t-SNE)^51^ to visualize the subsequence-level latent representations for females and males with AD. Fig. 2D shows the distribution of latent embeddings before (left panel) and after (right panel) training with LSTMAuto, which reveals a marked separation by sex, indicating that the model effectively captures sex differences in patients with AD. To identify the most suitable clustering algorithms for grouping sex-specific latent embeddings, we compared several classic clustering methods, i.e., k-means^52^, hierarchical agglomerative clustering^53^ (Agglomerative), spectral clustering^54^ (Spectral) and Gaussian mixture models (GMM)^55^. The evaluation was performed using three standard clustering indexes: silhouette score^56^ (range: [-1, 1], higher values indicate better-defined clusters), Davies–Bouldin index^57^ (range: [0, ∞), lower is better) and Dunn index^56^ (range: [0, ∞), higher is better). As shown in Fig. 2E, k-means achieved the best performance on the silhouette score (0.62) and the Davies– Bouldin index (0.51). Agglomerative clustering also showed competitive performance, indicated by the silhouette score (0.59) and the Dunn index (1.7e-03). Despite the marginally lower than K-means, we finally selected Agglomerative as our clustering method due to its greater flexibility in identifying non-spherical cluster structures and ability to preserve hierarchical relationships, key for interpreting progression pathways. The resulting clusters were subsequently used to define and analyze sex-specific AD sub-phenotypes.

### Clusters of sex-stratified temporal subsequences from patients with AD

#### The selection of cluster number and visualization

To determine the optimum clusters (i.e., disease states; see “Methods” section) of sex-stratified patients with AD, we first applied hierarchical agglomerative clustering to the subsequence-level latent embeddings. We explored different cluster numbers (k ranging from 2 to 10) and evaluated the results using two clustering quality metrics: Adjusted Rand Index (ARI) > 0.8 and silhouette score > 0.5. Based on these criteria, the optimal clustering was obtained at a dendrogram height of 20, resulting in seven distinct clusters, denoted as C1 through C7. Figure 3A shows a principal component analysis (PCA) biplot with clear separation among the identified clusters. Principal Component 1 (PC1) accounted for >99% of the variance and exhibited significant variation across the seven clusters (Fig. 3A), which may reflect strong correlations among phenotypic patterns in the cohort. To statistically validate the differences in PC1 distributions among clusters, we conducted a two-sided Mann–Whitney U-test (𝑝-value = 7.4 × 10⁻³⁷; Fig. 3B). Cluster C4 was positioned at the most negative end along Principal Component 1 (PC1) (−4.96 ± 0.57), while Cluster C7 was positioned at the most positive end (6.23 ± 0.64). According to Fig. 3C, we also found that the sex distribution differs across clusters. For example, Cluster C1 displayed a relatively balanced distribution (921 females and 704 males with AD). In contrast, Clusters C2, C3, and C4 mainly consisted of females (percentage = 57.7%, 90.1%, and 88.0%), whereas Clusters C5, C6, and C7 were male-dominant, comprising 73.0%, 96.7%, and 75.4% of males with AD, respectively. These patterns suggest distinct sex-specific trajectories in AD progression.

**Fig. 3:**
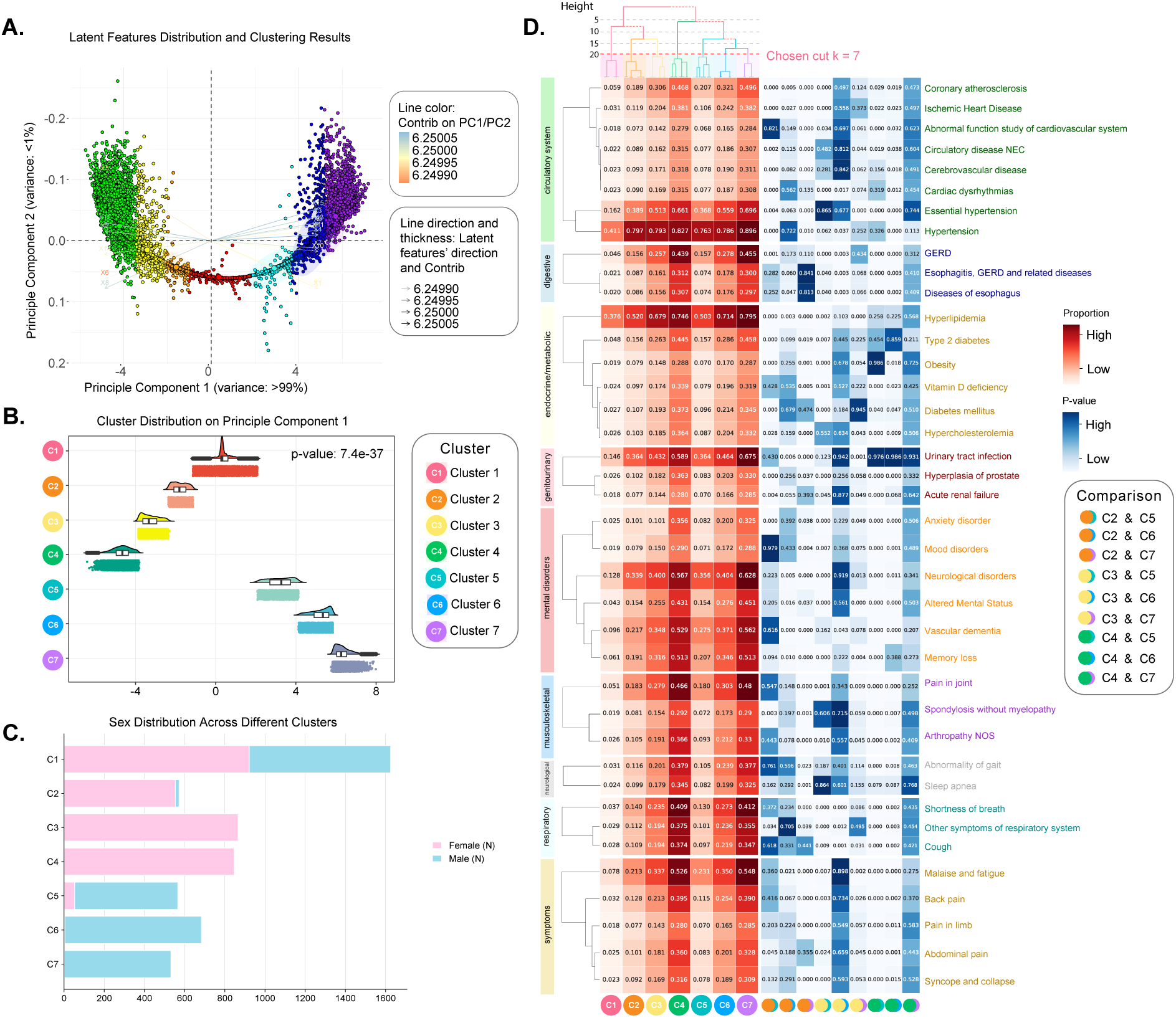
The characterization and analyses of derived clusters of sex-specific subsequences of patients with AD. **A.** The visualization of seven identified clusters (C1–C7, henceforth termed as states) using PCA. **B.** Violin plot for cluster distribution on the first principal component (The first principal component explained >99% of the variance in A.). **C.** The number of subsequences derived from females and males with AD in each cluster (i.e., state). **D.** The proportions of phenotypes in each state (left) and pairwise phenotypic comparisons between states (right). In the left panel, the intensity of the red color indicates the proportion of each phenotype within a state, while the accompanying dendrograms depict the hierarchical relationships among states and phenotypes. In the right panel, the depth of the blue color reflects the 𝑝-value from nine pairwise two-sided chi-square tests, with a significance threshold set at p < 0.05.

#### Phenotypic characteristics of distinct clusters

We examined the phenotypic profiles of the seven identified clusters (hereafter referred to as “states”) by visualizing their hierarchical relationships using dendrograms. As shown in Fig. 3D (upper-left), cutting the dendrogram at a height of 5 revealed two major groupings: clusters C1, C2, and C3 formed one branch, while C4 through C7 formed another, indicating structural distinctions between early and advanced disease states. To further characterize these clusters, we analyzed the distribution of phenotypes in patients with AD across nine clinical categories, including circulatory system, digestive, endocrine/metabolic, genitourinary, mental disorders, musculoskeletal, neurological, respiratory, and symptoms. These nine categories (selected from 18 Phecode Chapters; see “Methods” section) cover phenotypes that ranked in the top 50 in terms of phenotype prevalence proportion and showed statistically significant differences (i.e., 𝑝-value < 0.01) across all cluster-and subphenotype-level subgroups. The phenotype proportions within each cluster are displayed in Figure 3D (left) and detailed in Supplementary Data 1, highlighting the phenotypic variability and heterogeneity across disease states. We ranked the phenotypes based on their prevalence in cluster C1 and applied this ranking consistently across all clusters to enable direct comparison. In C1, we can see that the most prevalent conditions included hypertension (41.1%, circulatory system), hyperlipidemia (37.6%, endocrine/metabolic), essential hypertension (16.2%, circulatory system), urinary tract infection (14.6%, genitourinary), neurological disorders (12.8%, mental disorders), dementias (9.6%, mental disorders), malaise and fatigue (7.8%, symptoms), memory loss (6.1%, mental disorders), coronary atherosclerosis (5.9%, circulatory system), and pain in joint (5.1%, musculoskeletal). The results revealed that C4 and C7 exhibited a notable prevalence in multiple phenotypes across the nine categories. For example, within the circulatory system, hypertension was observed in 82.7% and 89.6% of patients in C4 and C7, respectively. In the digestive category, gastroesophageal reflux disease (GERD) affected 43.9% and 45.5% of patients in C4 and C7. Likewise, hyperlipidemia was present in 74.6% of C4 and 79.5% of C7 patients in the endocrine/metabolic category. In contrast, states C1 and C5 exhibit comparatively lower comorbidity burdens, with hypertension occurring in only 41.1% and 76.3% of patients in C1 and C5, respectively. These findings suggested that C4 and C7 represent disease states characterized by a high burden of comorbidities, while C1 and C5 correspond to disease states with relatively fewer comorbid conditions among patients with AD. Additional analysis revealed (Supplementary Table 3) that state C4 had the highest proportion of essential hypertension (82.7%) and the longest mean follow-up duration (8.32 years), followed by C3 (8.24 years), C2 (8.17 years), and C1 (8.1 years). This suggests that C4 may correspond to a more stable or slow progression pattern of the AD state, whereas patients in C1 might experience a more rapid disease progression. Similarly, we found that C5 exhibited both a lower comorbidity burden and a shorter mean follow-up duration (7.98 years) compared to C6 and C7, indicating a potentially distinct progression profile. Across all clusters, circulatory and endocrine/metabolic conditions, particularly hypertension, essential hypertension, and hyperlipidemia, emerged as the top 3 most prevalent comorbidities across AD disease states.

#### Analysis of phenotypic difference in pairwise clusters

To further assess phenotypic differences between states, we performed pairwise two-sided chi-square tests,^57^ measured by 𝑝-value, with a threshold 𝑝 < 0.05. The results (Fig. 3D, right) indicated significant differences regarding the top three prevalent phenotypes (i.e., hypertension, hyperlipidemia, and essential hypertension) in several cluster pairs, including C2 & C5 (𝑝-value < 1e-03, 𝑝-value < 1e-03, 𝑝-value = 4e-03) and C2 & C7 (𝑝-value = 1e-02, 0.000, 0.000). Other pairs, such as C3 & C5, C4 & C5, and C4 & C6, also showed significant differences in multiple phenotypes like coronary atherosclerosis and GERD, suggesting distinct comorbidity profiles across states, which is crucial to better understand the sex difference in AD progression. In contrast, clusters C4 and C7 displayed highly similar phenotypic distributions, with no significant differences across broader categories such as endocrine/metabolic, genitourinary, musculoskeletal, and symptom-related conditions. Additional non-significant comparisons included hypertension in C2 & C6 (𝑝-value = 0.722), hyperlipidemia in C4 & C5 (𝑝-value = 0.258), and essential hypertension in C3 vs. C5 (𝑝-value = 0.865). Moreover, we also found no significant differences in C3 & C6 and C4 & C7 pairs across broader phenotypic categories. These findings indicate that while most cluster pairs reflect meaningful phenotypic divergence, a few, notably C4 and C7, may represent closely related sub-phenotypes in AD progression.

### Sex-specific AD progression sub-phenotype identification

To characterize AD progression patterns, we constructed individual patient trajectories by temporally linking the identified disease states (i.e., clusters), which were used to derive sex-specific AD progression sub-phenotypes. Fig. 4A illustrates the example of six patients with AD with distinct progression pathways, represented by temporal clusters within the same patient along the timeline. For instance, the progression trajectory of the first female patient with AD (F1) was observed as a sequential transition through the states: C1→C1→C2→C3→C4→C4. Similarly, the AD progression of a male patient with AD (M2) was characterized as C1→C6→ C7→C7. We further quantified how frequently each disease state occurred within individual patient trajectories, measured by the number of times the same state appeared along a patient’s disease progression (Fig. 4B). The results suggest that Clusters C4 and C7 appeared most frequently and were sustained for longer durations compared to other clusters, such as C1, C2, and C3. These findings indicate that C4 and C7 represent more stable or enduring disease states in the progression pathways.

**Fig. 4:**
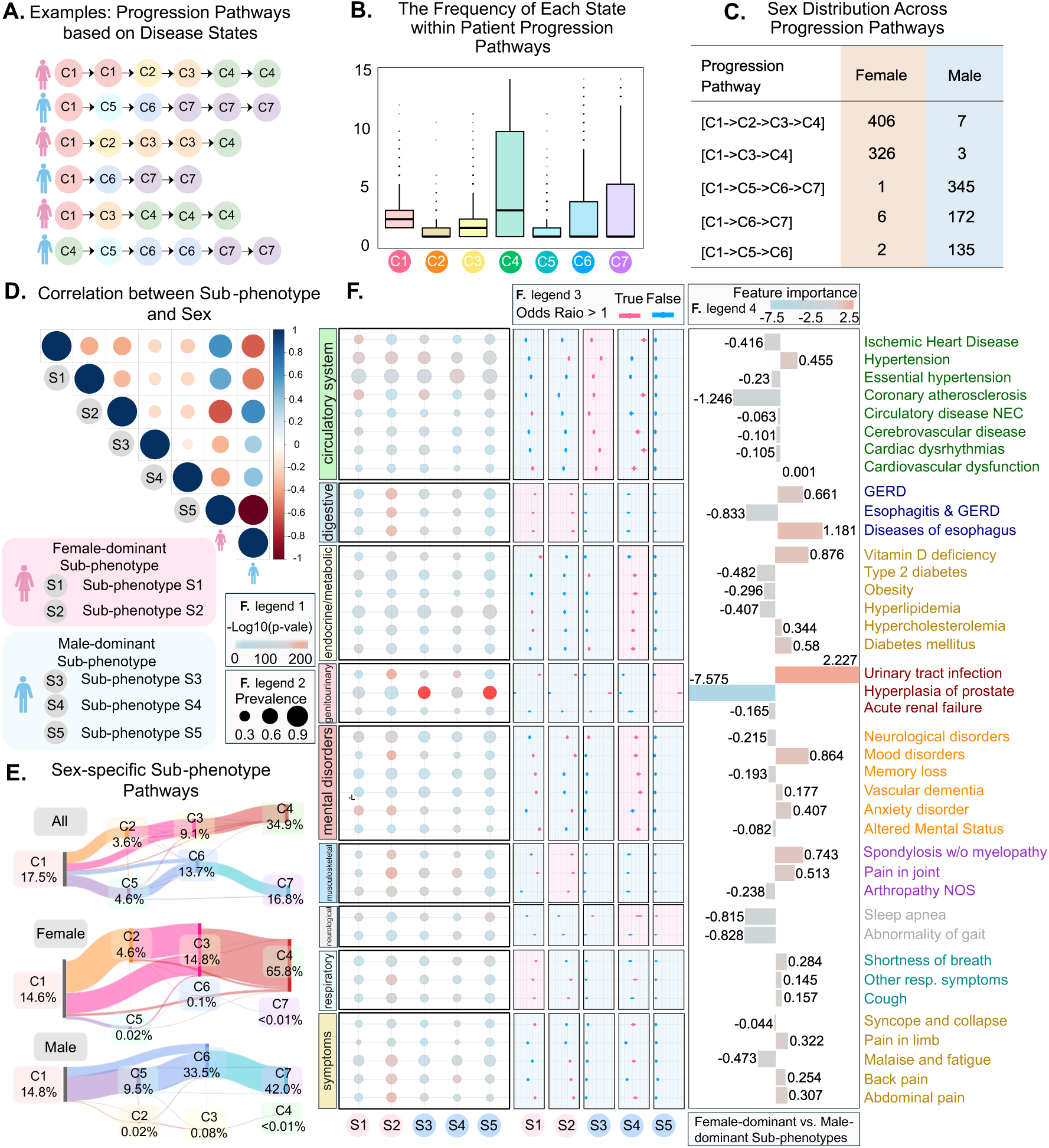
The derivation and characterization of sex-specific AD progression sub-phenotypes. **A.** The examples of the construction of patients’ progression pathways. **B.** The frequency of each disease state appeared within individual patient progression pathways. **C.** The number and progression pathways of the five primary identified sex-specific AD sub-phenotypes. **D.** The correlation between AD sub-phenotype and sex. **E.** The Sankey diagrams of AD progression pathways for all patients (top), females with AD (middle), and males with AD (bottom). **F.** The phenotype-based analyses of the five sex-specific AD sub-phenotypes: prevalence, statistical significance, and predictive importance. The bubble maps (left) display the proportions and statistical significance of phenotypes within each sub-phenotype, where bubble size corresponds to -log10 (𝑝-value). The forest plot (middle) presents the odds ratios (ORs) of phenotype proportions across sub-phenotypes, with red markers indicating ORs greater than 1. The red background highlights phenotypes particularly associated with their respective sub-phenotypes. The pyramid plot (right) illustrates feature importance for classifying female- and male-dominant sub-phenotypes, with bar length representing the magnitude and direction of each phenotype’s contribution.

We then consolidated consecutive identical states (e.g., C1→C6→C7→C7 to C1→C6→C7) to generate a refined progression pathway for each patient with AD. Subsequently, we counted the number of patients sharing the same refined pathways, and identified five primary sex-specific progression patterns, each included at least 100 patients (Fig. 4C) and where each state in the trajectory accounted for more than 1% of the total sub-phenotype population (Fig. 4E). These sub-phenotypes, denoted as S1-S5, represent distinct progression pathways, where (1) S1: C1→C2→C3→C4, (2) S2: C1→C3→C4, (3) S3: C1→C5→C6→C7, (4) S4: C1→C6→C7, and (5) S5: C1→C5→C6. According to Fig. 4C, sub-phenotypes S1 and S2 were predominantly composed of females with AD, with 406 and 326 cases, but only 7 and 3 males, respectively. In contrast, S3, S4, and S5 are male-dominant AD sub-phenotypes, consisting of 345 vs 1, 172 vs 6, and 135 vs 2 for male and female cases. Additionally, correlation analysis further confirmed that females with AD showed a stronger correlation with S1 and S2, whereas males with AD were closely associated with sub-phenotypes S3, S4, and S5 (Fig. 4D). These associations are visually reflected in both color and size, where color indicates the direction of correlation (blue for positive, red for negative), and circle size represents the magnitude of correlation (Fig. 4D). We also used Sankey diagrams to illustrate the progression pathways and transitions across all, females and males with AD, respectively (Fig. 4E). The width of the Sankey diagram represents the size of patients transiting from one state to another. The percentage values under each cluster indicate the overall proportion of patients who passed through that state in their disease trajectory, highlighting which states were most commonly encountered in the disease progression across the population. Interestingly, we noticed that in the flow, the states C2, C3, and C4 only existed in female-dominant sub-phenotypes S1 and S2, whereas C5, C6, and C7 were uniquely present in male-dominant sub-phenotypes S3, S4, and S5. The results further validated different progression patterns in females and males with AD with distinct sub-phenotypes.

### Phenotypic characteristics of sex-specific sub-phenotypes

We conducted statistical analyses on the five identified sub-phenotypes, with results summarized in Table 2. We observed that sub-phenotype S1 had the highest mean age at AD diagnosis (mean age: 77.2 ± 9.8 years), the longest follow-up duration (8.31 years), and the longest disease duration (2.64 years). In contrast, sub-phenotype S5 exhibited the lowest mean age at AD diagnosis (mean age: 74.8 ± 9.5 years), along with the shortest follow-up (7.39 years) and disease duration (2.14 years). In terms of racial composition, S5 had the highest proportion of Hispanic individuals (n = 38, 27.7%), while S1 had the largest number of Hispanic individuals (n = 111, 26.9%). Sub-phenotype S2 showed a relatively higher percentage of black or African American individuals (n = 60, 18.2%) compared to other sub-phenotypes, whereas S3 had a higher percentage of White individuals (n = 279, 80.6%). Mortality also varied across sub-phenotypes, with S3 exhibiting the highest rate (15.9%) and S5 the lowest (7.3%). In terms of the three most common comorbidities of AD, neurological disorders were most prevalent in sub-phenotype S3 (41.3%), followed by S4 (39.3%) and S1 (36.1%). Meanwhile, cardiovascular diseases also showed the highest prevalence in S3 (22.5%), followed by S2 and S4 (19.8% and 19.1%, respectively). Diabetes-related conditions were most prevalent among patients with AD in sub-phenotype S4 (51.7%). More demographic and phenotypic characteristics for each sub-phenotype are provided in Table 2 and Supplementary Table 4.

**Table 2.** Descriptive statistics of five identified primary sex-specific AD sub-phenotypes.

| Characteristics | S1<br>(N = 413,<br>24.8%) | S2<br>(N = 329,<br>19.8%) | S3<br>(N = 346,<br>20.8%) | S4<br>(N = 178,<br>10.7%) | S5<br>(N = 137,<br>8.2%) |
| --- | --- | --- | --- | --- | --- |
| **Demographics** |  |  |  |  |  |
| Age at AD diagnosis, mean (std) | 77.2 (9.8) | 76.9 (9.2) | 76.2 (8.7) | 77.1 (8.3) | 74.8 (9.5) |
| Female, N (%) | 406 (98.3%) | 326 (99.1%) | 1 (0.3%) | 6 (3.4%) | 2 (1.5%) |
| Male, N (%) | 7 (1.7%) | 3 (0.9%) | 345 (99.7%) | 172 (96.6%) | 135 (98.5%) |
| **Disease development, mean years (std)** |  |  |  |  |  |
| Follow-up durations | 8.31 (1.61) | 8.29 (1.7) | 8.24 (1.57) | 8.05 (1.67) | 7.39 (1.80) |
| Years before diagnosis | 5.67 (1.73) | 5.69 (1.76) | 5.76 (1.66) | 5.68 (1.80) | 5.25 (1.52) |
| Years after diagnosis | 2.64 (1.56) | 2.6 (1.54) | 2.48 (1.46) | 2.37 (1.52) | 2.14 (1.42) |
| **Hispanic, N (%)** |  |  |  |  |  |
| Hispanic | 111 (26.9%) | 84 (25.5%) | 85 (24.6%) | 43 (24.2%) | 38 (27.7%) |
| Not Hispanic | 293 (70.9%) | 240 (72.9%) | 260 (75.1%) | 128 (71.9%) | 97 (70.8%) |
| No Hispanic information | 2 (0.5%) | 2 (0.6%) | 1 (0.3%) | 1 (0.6%) | 1 (0.7%) |
| **Race, N (%)** |  |  |  |  |  |
| American Indian or Alaska Native | 0 (0.0%) | 0 (0.0%) | 0 (0.0%) | 0 (0.0%) | 0 (0.0%) |
| Asian | 3 (0.7%) | 0 (0.0%) | 3 (0.9%) | 1 (0.6%) | 1 (0.7%) |
| Black or African American | 56 (13.6%) | 60 (18.2%) | 55 (15.9%) | 26 (14.6%) | 12 (8.8%) |
| White | 321 (77.7%) | 245 (74.5%) | 279 (80.6%) | 133 (74.7%) | 109 (79.6%) |
| Multiple race | 2 (0.5%) | 2 (0.6%) | 3 (0.9%) | 2 (1.1%) | 3 (2.2%) |
| Unknown | 24 (5.8%) | 17 (5.2%) | 6 (1.7%) | 10 (5.6%) | 11 (8.0%) |
| **Comorbidity, N (%)** |  |  |  |  |  |
| Neurological | 149 (36.1%) | 113 (34.3%) | 143 (41.3%) | 70 (39.3%) | 37 (27.0%) |
| Cardiovascular | 68 (16.5%) | 65 (19.8%) | 78 (22.5%) | 34 (19.1%) | 21 (15.3%) |
| Diabetes-related | 171 (41.4%) | 157 (47.7%) | 178 (51.4%) | 92 (51.7%) | 50 (36.5%) |

To characterize the phenotypic characteristics of the identified sex-specific sub-phenotypes of patients with AD, we investigated the patient percentages based on the top phenotypic features in nine categories (Fig. 4F, left; Supplementary Data 2). Among these, sub-phenotype S4 had the largest proportion of essential hypertension at 79.1%, a condition also prevalent in other sub-phenotypes, such as S1 (78.7%), S5 (77.9%), and S2 (75.3%). This finding suggests that essential hypertension is a key phenotype shared by these patients with AD, regardless of sex. Similarly, hyperlipidemia and hypertension were frequently observed in both female- and male-dominant sub-phenotypes, aligning with the top three phenotypes previously identified in individual disease states. Among male-dominant sub-phenotypes, aside from the top three phenotypes, hyperplasia of prostate, as a male-specific phenotype, was also notably prevalent (63.8%). Additionally, S5 showed substantially higher proportions in most phenotypes, whereas S4 consistently exhibited lower proportions. In contrast, female-dominant sub-phenotype S2 showed elevated proportions of neurological disorders (49.9%) and malaise and fatigue (45.8%). Moreover, we performed pairwise chi-square tests to investigate the differences in clinical phenotypes across sex-specific AD sub-phenotypes. In addition to commonly prevalent phenotypes, several other conditions showed strong sub-phenotype-specific associations. For example, hyperplasia of prostate was significantly enriched in male-dominant sub-phenotypes S3 (-log₁₀ *p =* 2.89e+02, 95% CI: 15.1,18.1) and S5 (-log₁₀ *p =* 2.89e+02, 95% CI: 3.62, 4.19). Coronary atherosclerosis was particularly associated with S3 (-log₁₀ *p =* 7.19e+01, 95% CI: 1.68, 1.91), whereas sleep apnea showed a significant association in S5 (-log₁₀ *p =* 5.20e+01, 95% CI: 1.89, 2.27). Conversely, the female-dominant sub-phenotype S2 was significantly associated with multiple phenotypes, including urinary tract infection (-log₁₀ *p =* 1.39e+02, 95% CI: 2.15, 2.45), mood disorders (-log₁₀ *p =* 1.28e+02, 95% CI: 2.09, 2.38), and pain in joint (-log₁₀ *p =* 1.181e+02, 95% CI: 1.95, 2.21). These findings highlight distinct clinical profiles within sex-stratified AD sub-phenotypes.

To further investigate unique phenotypic features within sex-specific AD sub-phenotypes, we focused on phenotypes with pooled odds ratios larger than 1, highlighting those with stronger associations (Fig. 4E middle, with red indicating greater significance). In the female-dominant sub-phenotypes, both S1 and S2 showed elevated effect sizes in digestive disorders, including gastroesophageal reflux disease (GERD), esophagitis, and other esophageal diseases. Additionally, S1 was associated with increased effect sizes for respiratory symptoms like shortness of breath, cough, and other respiratory issues, while S2 exhibited stronger associations with musculoskeletal disorders such as spondylosis, joint pain, and arthropathy not otherwise specified. Among the male-dominant sub-phenotypes, S3, S4, and S5 demonstrated higher effect sizes for neurological conditions, particularly sleep apnea. S3 and S5 were also highly associated with genitourinary disorders (e.g., hyperplasia of the prostate), while S4 was enriched for endocrine/metabolic conditions (e.g., type 2 diabetes, obesity, hyperlipidemia, hypercholesterolemia, diabetes mellitus) and mental disorders (e.g., neurological disorders, mood disorders, memory loss and vascular dementia). To assess the discriminative power of identified sex-specific AD sub-phenotypes, we combined female-dominant (S1, S2) and male-dominant (S3–S5) sub-phenotypes as groups including S1 & S3, S1 & S4, S1 & S5, S2 & S3, S2 & S4, and S2 & S5. We trained the regression models to classify these groups based on phenotypic features. The average model coefficients were used to infer phenotype importance (Fig. 4F, right), and we identified several top phenotypic predictors. It is shown that hyperplasia of prostate (genitourinary, coefficient = -7.575) was the most prominent predictor for male-dominant sub-phenotype, while urinary tract infection (genitourinary, coefficient = 2.227) was strongly associated with female-dominant sub-phenotype. Other top predictors for the male-dominant group included coronary atherosclerosis (circulatory system, coefficient = -0.828) and abnormal gait (neurological, coefficient = -0.828). In contrast, the key predictors for female-dominant groups consisted of diseases of the esophagus (digestive, coefficient = 1.181) and mood disorders (mental disorders, coefficient = 0.864), suggesting differing symptom burdens and comorbidity profiles between sex-based AD groups.

### Analysis of AD progression patterns in sex-specific sub-phenotypes

To gain deeper insights into how progression patterns differ across sex-specific sub-phenotypes, we conducted a two-fold analysis, i.e., survival analysis using the Kaplan-Meier method^58^ and Cox proportional hazards analysis^59^. The results are presented in Fig. 5A, and we adjusted for the three most prevalent comorbid conditions, i.e., essential hypertension, hypertension, and hyperlipidemia, as observational variables, using AD diagnosis as the outcome event. The analysis was performed over a 5-year observation window, measuring survival time from the index visit to the first documented AD diagnosis. In Fig. 5A (top), the x-axis represents time in days, and the y-axis shows the fraction of patients who have not yet received an AD diagnosis (i.e., survival probability without AD). Our findings revealed that the male-dominant sub-phenotype S5 had the fastest decline in survival probability without AD compared to other sub-phenotypes when either essential hypertension or hyperlipidemia was exposed within patients. In contrast, female-dominant sub-phenotype S2 has the fastest decline in survival probability for those who are under hypertension. Furthermore, S1 (female-dominant) and S4 (male-dominant) demonstrated the slowest decline in survival probability across all three conditions. The results also showed the temporal changes over time in the number of patients who had not yet been diagnosed with AD (i.e., still at risk of AD) across sex-specific sub-phenotypes (Fig. 5A, bottom). For example, under essential hypertension, both female-and male-dominant sub-phenotypes showed a faster decline in the number of patients’ survival without AD over time, indicating a quicker loss of AD-free states. It was followed by hyperlipidemia, while hypertension showed the slowest decline. These results, along with the two-sided log-rank tests (Log-rank 𝑝 -value < 0.0001), indicate significant differences in survival probabilities across sub-phenotypes under different comorbidities. The difference in the rate of decline in survival probabilities without AD between hypertension and essential hypertension may be due to the broader range of related diagnoses included under the hypertension phenotype, which may cover milder or incidental cases. This highlights the importance of using more specific comorbid phenotypes in analysis. Cox proportional hazards analysis further confirmed that both essential hypertension and hypertension were significantly associated with increased AD risk. The hazard ratios (HRs) under all three comorbidity conditions were greater than 1, but only essential hypertension and hypertension had confidence intervals that did not include 1 (essential hypertension: HR = 1.6 [95% CI: 1.05, 2.43]; hypertension: HR = 1.81 [95% CI: 1.22, 2.70]; hyperlipidemia: HR = 1.4 [95% CI: 0.901, 2.243]). These findings suggest that essential hypertension and hypertension significantly increase the risk of developing AD. They also highlight the substantial heterogeneity in AD progression across sex and comorbidity conditions, underscoring the importance of stratifying patients by sex-specific sub-phenotypes.

**Fig. 5:**
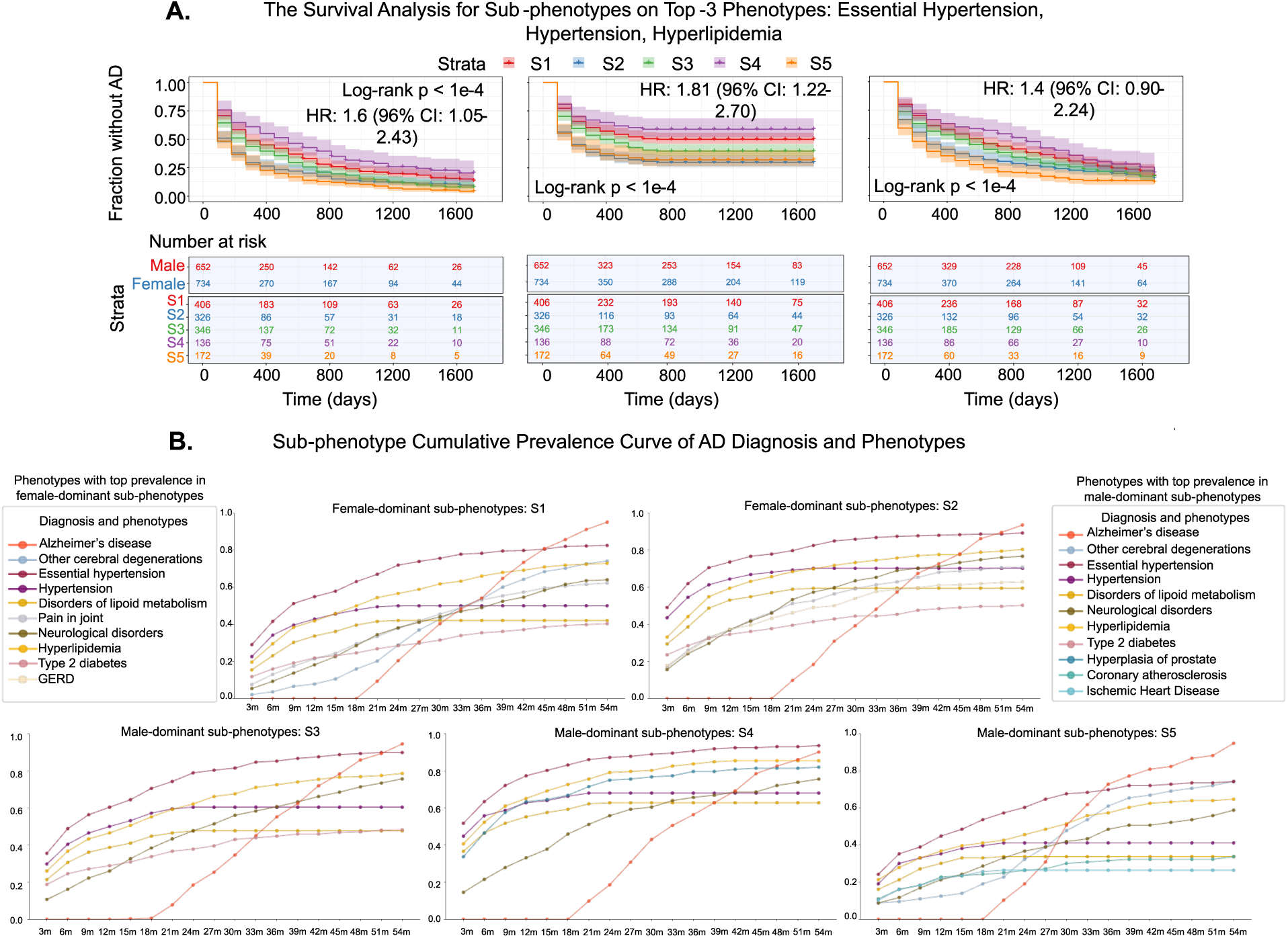
Survival analysis and cumulative prevalence curves for top comorbid phenotypes associated with AD onset across five identified sex-specific sub-phenotypes. **A.** Kaplan-Meier survival curve depicting time to AD onset across the top three comorbid phenotypes for each sub-phenotype. The curves represent the proportion of patients without AD over time, with the Log-rank test’s p-values indicating significant differences between sub-phenotypes (p < 1e-4). The number of patients at risk is displayed for each sub-phenotype at different time points (unit: day). **B.** Cumulative prevalence curves for the five sex-specific AD sub-phenotypes, showing the accumulation of high-prevalence comorbidities or AD onset over time. Each point represents the cumulative proportion of patients diagnosed with either AD or one of the most prevalent comorbid phenotypes, calculated at 3-month intervals.

We also calculated the cumulative prevalence of selected comorbidity phenotypes (i.e., essential hypertension, hyperlipidemia, hypertension, and neurological disorders) along with AD prevalence across different sub-phenotypes over time. The observation window was 54 months, divided into 3-month intervals to track cumulative prevalence. As shown in Fig. 5B, AD prevalence across all sub-phenotypes began to increase around the 18^th^ month and continued to rise until the 54^th^ month. Among the selected comorbid phenotypes, essential hypertension showed the highest cumulative prevalence, followed by hyperlipidemia and hypertension. This finding is consistent with our statistical analysis, which identified these three phenotypes as having the highest prevalence across the cohort. At the sub-phenotypes level, we found that S2 and S4 exhibited the earliest accumulation of comorbidities, with each exceeding 30% prevalence within the first 3 months. The cumulative prevalence of AD eventually exceeded comorbidities at different time points across sub-phenotypes. For example, in S1, AD prevalence surpassed hypertension between Month 33–36 and all three comorbidities between months 45–48. In S2, this crossover occurred between Month 39–42 for hypertension and Month 48–51 for all these three comorbidities. Similar patterns can also be observed in sub-phenotypes S3 and S4. Notably, S5 displayed the fastest disease progression, with AD prevalence exceeding hypertension as early as Month 27–30 and all top comorbidities in Month 33–36. Aligned with the survival analysis, S5 showed both the earliest and steepest increase in AD prevalence, along with a faster decline in survival probability, indicating a need for earlier clinical intervention during the initial phase of comorbidity accumulation. In contrast, although S1 also showed a relatively rapid increase in AD prevalence, it maintained a slower decline in survival probability, while S3 followed a relatively stable progression trajectory but was linked to the male-specific comorbidity hyperplasia of prostate. These findings highlight the temporal and sex-specific heterogeneity in AD progression and suggest that monitoring comorbidity trends could be an effective strategy for predicting AD risk and informing early intervention strategies across different sub-phenotypes.

## Discussion

In this work, we developed a novel auto-encoder framework to identify sex-specific sub-phenotypes of AD progression using longitudinal EHRs and uncover their associated clinical characteristics. The OneFlorida+ Clinical Research Consortium enabled us to collect large-scale and heterogeneous clinical information from patients with AD, including diagnosis, lab measurements, medication, and more. We then constructed temporal subsequences from each patient’s record to capture their disease progression trajectory. Our approach integrated a deep learning-based autoencoder with clustering techniques to derive meaningful sex-specific AD sub-phenotypes. The autoencoder generated sex-based embeddings of temporal subsequences representing various clinical information of patients with AD. We then applied clustering algorithms to group these embeddings into different clusters (i.e., disease states). By concatenating the states corresponding to each patient’s subsequences, we constructed individual progression trajectories, or sub-phenotypes, from which we extracted characteristic clinical patterns. We evaluated our framework by AUROC and clustering robustness (e.g., silhouette score), confirming the stability and reliability of our model. Ultimately, we uncovered five primary sex-specific AD sub-phenotypes with distinct clinical characteristics (Fig. 4C). Our findings highlight the heterogeneity of AD progression and underscore the critical role of incorporating sex-specific factors into disease modeling and personalized treatment strategies. The identified sub-phenotypes not only represent divergent progression pathways but also illuminate sex-based differences in clinical characteristics and comorbidities of patients with AD. In our cohort, females showed a larger proportion of patients with AD, with an older age of diagnosis and a longer time of disease development on average. However, males with AD had a higher mortality rate (14.20%) compared to females (11.34%), consistent with findings from previous studies^1,40,60^. Through our clustering analysis, we identified seven distinct disease states (C1–C7), with state C4 encompassing the largest number of patient subsequences. Examination of clinical characteristics across these states revealed that essential hypertension (41.1%) was the most prevalent phenotype, followed by hyperlipidemia (37.6%), hypertension (16.2%), hyperplasia of prostate (14.6%), and neurological disorders (12.8%). Although previous studies have established associations between these comorbidities and sex-specific AD pathology, our study further revealed, through pairwise comparisons using chi-square tests, that these comorbidities exhibited similarities between some disease states within sex-specific progression pathways, while showing significant differences between some other disease states. This provided new insights into the characterization of AD disease states. For example, certain states (e.g., C4 and C7) shared similar phenotype proportions across categories such as endocrine/metabolic and genitourinary systems. Some states (e.g., C2 vs. C5, C2 vs. C7) showed statistically significant differences in phenotypes like hypertension, hyperlipidemia, and essential hypertension. Moreover, these disease states demonstrated different sex distributions. These findings suggested that sex not only influences the overall clinical profiles of AD but also contributes to the homogeneity or heterogeneity of particular disease states and their associated comorbidities, emphasizing the importance of identifying sex-specific progression pathways composed of distinct disease states.

Based on the aggregation of disease states following temporal orders, we constructed patients’ disease progression pathways and identified five major AD sub-phenotypes. Among them, two were female-dominant (S1 and S2), and the remaining three were male-dominant (S3, S4, and S5), each exhibiting distinct progression pathways (Fig. 4C). Analysis of phenotype proportions suggested that, among all sex-specific AD sub-phenotypes, comorbidities with the highest prevalence were circulatory system disorders (i.e., essential hypertension and hypertension) and endocrine/metabolic disorders (i.e., hyperlipidemia). These comorbid phenotypes have also been widely recognized in prior studies as risk factors in AD progression. The high prevalence and average statistical significance level of hypertension and other vascular conditions support existing hypotheses that vascular disease^61–64^ and neuroinflammation^65^ may contribute to AD progression. These comorbidities may either directly drive neurodegeneration or expose AD symptoms by reducing cerebrovascular reserve and cognitive resilience through vascular brain injury. In addition, we characterized the clinical profiles of each sub-phenotype. Based on the prevalence proportion among phenotypes, we observed that sub-phenotype S1 had a lower overall comorbidity burden compared to S2. By calculating the pooled odds ratios from pairwise chi-square tests among sub-phenotypes, we also identified several distinct phenotypes for S1, characterized by a higher prevalence of respiratory conditions, including shortness of breath, cough, and other respiratory symptoms. While respiratory diseases are not typically considered directly associated with AD, our findings suggest that they influence disease progression among some specific females with AD. According to previous research, they influence AD progression possibly through indirect mechanisms such as disrupted sleep patterns^66^, increased exposure to air pollution^67^, or susceptibility to pulmonary infections^68^ in elderly patients with AD. Compared to S1, sub-phenotype S2 included a higher proportion of Black or African American individuals and demonstrated broader comorbidity prevalence across multiple phenotype categories, indicating a greater comorbidity burden. For S2, neurological disorders were a primary phenotype, supporting previous studies suggesting that women with AD have a stronger association with mental disorders than men, such as depression^69^ and mood disorders^70^. For male-dominant sub-phenotypes, S3 had the largest number of males and the highest proportion of White individuals. This subgroup exhibited statistically significant disorders affecting the circulatory and genitourinary systems. Sub-phenotype S4 presented the lowest overall comorbidity burden within the male sub-phenotypes, and it was notably characterized by a concentration of endocrine and metabolic disorders, including diabetes, obesity, hypercholesterolemia and other forms of diabetes mellitus. In contrast, S5 demonstrated the highest comorbidity burden among the male-dominant sub-phenotypes, characterized by several significant phenotypes, particularly genitourinary disorders (e.g., hyperplasia of prostate) and neurological conditions (e.g., sleep apnea and abnormality of gait). Moreover, it was notable that in our findings, both females and males with AD exhibited genitourinary system disorders, specifically hyperplasia of the prostate in males and urinary tract infection (UTI) in females. It appeared to support the growing hypothesis that genitourinary issues are common among older adults and increasingly recognized as being associated with AD. In particular, UTI in women have been linked to systemic inflammation and delirium, which may exacerbate cognitive decline in patients with AD^71^. Interestingly, our analysis revealed some novel associations beyond the commonly recognized AD-related phenotypes. Specifically, females with AD had a strong association with digestive disorders, including GERD, esophagitis, GERD-related diseases, and diseases of the esophagus, areas that have received limited attention in AD research. Although emerging evidence suggests an association between gastrointestinal disorders and both central and peripheral nervous system diseases^72^, sex differences in these associations remain limited explored.

We further performed progression analysis and observed that the identified sub-phenotypes exhibited distinct disease dynamics along their progression pathways. For example, compared with S3 (C1 → C5 → C6 → C7), the trajectory of S4 (C1 → C6 → C7) lacks the intermediate state C5, suggesting a potentially more direct transition through advanced disease stages. Survival analysis revealed that comorbidity burden influences the rate of AD progression differently across sub-phenotypes. The decline rates in survival probabilities prior to AD onset varied according to exposure to the three most prevalent comorbid conditions. Among patients with essential hypertension, the male-dominant sub-phenotype S5 presented the fastest progression to AD, followed by S2, a female-dominant sub-phenotype, both associated with higher comorbidity burdens. In contrast, S1 (female-dominant) and S4 (male-dominant) showed the slower decline of survival probabilities and were characterized by relatively lower comorbidity burden. These findings suggest that a greater comorbidity burden is potentially linked to accelerated AD onset. In patients with hypertension, S2 demonstrated the most rapid progression, consistent with previous reports that postmenopausal hypertension and depressive symptoms may synergistically exacerbate dementia progression in women^73^. Our results further supported the role of vascular and psychiatric risk factors in sex-specific AD pathogenesis. Moreover, cumulative prevalence analysis confirmed that S5 experienced the steepest and earliest rise in AD prevalence, emphasizing the need for early intervention, even during the initial stages of comorbidity development. By contrast, S4 exhibited a comparatively slower accumulation of AD prevalence over time. Collectively, these underscore the importance of examining both additive and interactive effects of comorbidities on AD progression in a sex-specific context, highlighting opportunities for targeted early interventions.

This study, while insightful in exploring and identifying sex differences of AD sub-phenotypes through advanced AI techniques, has several limitations that should be acknowledged. First, the reliance on EHR from one specific research network may introduce biases due to the demographic and geographic characteristics of the population. The patients in OneFlorida+, predominantly from Florida, Georgia, and Alabama, might not fully represent the broader, more diverse U.S. population or other global populations. The generalizability of the findings should be further validated with heterogeneous demographic profiles. Second, while the clustering model showed stability and reproducibility, the interpretation of the sex-specific AD sub-phenotypes remains challenging. The clinical significance of these clusters, especially their specific meaning and potential utility in guiding personalized treatment plans, needs further validation through prospective studies and clinical trials. The study’s design limits the ability to identify detailed causal inferences about the progression pathways and their implications for disease development. Third, although we discovered several sex-specific sub-phenotypes using data-driven methods, this study primarily relies on routine EHR data, lacking detailed cognitive assessment (e.g., the Mini-Mental State Exam) and biomarker data (e.g., brain imaging or cerebrospinal fluid analysis), which are crucial in AD progression and sex difference study. The incorporation of these data categories should be considered in future investigations, which can provide a more comprehensive and valuable insight into the potential mechanisms driving sex differences in AD progression and develop personalized therapeutic interventions.

Future research could prioritize three key directions upon the findings of this study. First, there is a critical need to validate the identified sub-phenotypes in larger and more demographically diverse populations to improve the generalizability and clinical applicability of the findings. This underscores the need to adopt methods such as federated learning techniques^74^ that can scale across large heterogeneous datasets. Second, further investigation into the biological mechanisms underlying sex differences in AD progression is essential for the development of sex-stratified therapeutic interventions. This highlights the necessity of employing multimodal and multi-omics data and research approaches^75^. For example, incorporating the genetic data of females and males with AD through whole genome or exome sequencing analyses can identify potential differential genes and variants associated with clinical manifestation in AD development. Thirdly, developing more robust and powerful computational models to precisely and quantitatively trace and identify phenotypic changes in the development of AD for individuals, which can provide timely monitoring and feedback for disease management.

## Methods

### Data source and cohort identification

The large amount of longitudinal EHR data of patients with AD was derived from the OneFlorida+ Clinical Research Consortium. As of 2025, OneFlorida+ covers more than 26 million patients across regions, including Florida, Atlanta (Georgia), Birmingham (Alabama), Arkansas, Minnesota, and Irvine (California), through its collaborative network of 14 health institutions comprising community health systems, clinics, and academic medical centers^76–78^. It consists of diverse EHR-based information adhering to the PCORnet Common Data Model (CDM), including demographics, encounters, procedures, diagnoses, vital signs, lab results, and others. To create the study cohort, we randomly selected ten sites, specifically sources 1, 3, 6, 9, and sources 10-15. This study complied with all relevant ethical regulations and was approved by the University of Florida Institutional Review Board (protocol no. IRB202202820). All patients’ clinical data were de-identified, and the institutions waived the requirement for written informed permission. We identified the patient cohort by the following criteria: (1) The start of the observation time was defined as the earliest of January 2012; (2) The first visit should be at least 50 years of age or older; (3) Patients have at least one diagnosis code of AD, i.e., International Classification of Diseases, Ninth Revision, Clinical Modification (ICD-9-CM) codes: 331.0 and International Classification of Diseases, Tenth Revision, Clinical Modification (ICD-10-CM) codes G30.0, G30.1, G30.8, and G30.9^42^. (4) The patients have at least two encounters. (5) Patients have at least one visit record three years before AD diagnosis and over one year of disease progression after AD diagnosis. We excluded the patients with AD diagnosis dates that are not within the period between their earliest and latest visit times.

### Variable selection and preprocessing

The EHR information from selected patients with AD consists of thousands of features with various structures and multiple types. To better preprocess, we categorize these features into four structural types, i.e., continuous features (e.g., body mass index and age), binary features (e.g., death), time-to-event features (e.g., diagnosis and treatment), and multi-class features (e.g., smoking status and medication code). These features were transformed with different strategies based on categories to ensure they can be processed by the classifiers. For instance, age at index was discretized using bins of identical 10-year size, and BMI was discretized into four groups: underweight (≤18.5), normal weight (18.5–24.9), overweight (25–29.9), and obesity (≥30) (excluding BMI values of 0 and those over 100 as outliers). In addition to variability in data structures, each clinical data source, including demographics, diagnoses, procedures, vital signs, medications, and lab results, used distinct coding systems. For the sake of standardization, we applied consistent clinical code mapping for each data type. For diagnosis, the codes were mapped to Phecodes, an EHR-specific codebase that supports phenome-wide association studies (PheWAS^79^. Since clinical diagnosis is based on clinical phenotypes that differ across AD stages^80^, we grouped these Phecodes into a variable set, renamed as phenotypes. For medication, we mapped National Drug Codes (NDC) and RxNorm codes into the unified Anatomical Therapeutic Chemical (ATC) Classification codes^81^. In terms of lab results, using Logical Observation Identifiers Names and Codes (LOINC) and Current Procedural Terminology (CPT), all LOINC codes were mapped to CPT^82^. Further, we applied Multiple Imputations by Chained Equations (MICE) to tackle the missingness in all variables^83^. All these variables were one-hot encoded, and finally, we concatenated them into a patient-centered embedding, excluding sex, which was used as the prediction label.

### Constructing temporal patient representation using longitudinal EHRs

To construct patients’ longitudinal progression trajectory covering the three years before and one year after AD onset, we first aggregated multiple measurements or events recorded at irregular time points and converted them into fixed time-interval blocks (e.g., 3-month time intervals) to form subsequences for each patient. Each block formed a vector representing a specific event type (e.g., diagnoses, medications, etc.). To model the progression patterns, we split each patient into multiple temporal sub-sequences. Fig. 1B and Supplementary Fig. 1 present the construction of the AD temporal trajectory using longitudinal EHRs. For example, a diagnosis vector contained distinct diagnosis codes and their frequencies within a 3-month window. For the invariant variables, demographic (e.g., race and gender), we treated them as static features and passed them at each time window. To define the longitudinal observation window for each patient, we selected the earliest encounter date within three years prior to AD onset as the start date (𝑇*_*N*_*), the first recorded AD diagnosis as the diagnosis date (𝑇_*diag*_), and the latest encounter after AD onset as the end date (𝑇_*m*_). Given 𝑁 patients, the total number of time windows 𝑇, and the dimension of patient feature matrix 𝐷, we constructed the temporal patient representation. The representation is a matrix in which patients can be represented as a 3-tuple symbol and a sequence of vectors for patient 𝑖 at visit 𝑡_*j*_, where 𝑡_*j*_ ∈ {𝑡*_1_*, 𝑡*_2_*, …, 𝑡_*T*_} and each visit is denoted as 𝑥_*i,t_j_*_ ∈ ℝ^*T×D*^. To increase the quantity and augment the data, the original data (where each patient corresponds to a temporal matrix) has been divided into multiple subsequences. Starting from the start date 𝑡*_1_* (the date of the first visit), every 2 windows (e.g., 6 months) form one subsequence, until reaching the maximum length of the patient’s EHR data. We then divided each patient into multiple subsequences with varying time lengths, starting from the index date, and new subsequences are created with 3-month increments (i.e., 6-month, 9-month, etc.) until the last encounter of patients within the end date. Each subsequence was treated as an independent temporal matrix sample and input into the model. Therefore, the 𝑙-th subsequence can be represented as 𝑥_*i,t_j_*_^(*l_j_*)^, where {𝑙*_1_*, 𝑙*_2_*, …, 𝑙_*N*_} is the index of each patient’s subsequence. Finally, we obtained the entire temporal matrix with dimensions from (𝑁, 𝑇, 𝐷) to (∑_*n*=1_^*N*^ 𝑙_*n*_, 𝑇, 𝐷). We concatenated these subsequences in different time lengths to construct the AD progression trajectory for progression modeling.

### Generating sex-specific AD latent representation with autoencoder

To extract latent embeddings of the subsequences of patients with AD, we employed an autoencoder architecture, consisting of an encoder-decoder structure, which compressed high-dimensional temporal EHRs of patients with AD into a lower-dimensional latent space, preserving critical temporal information. We first constructed a machine learning-based autoencoder, comprising a three-layer fully connected encoder and a symmetric three-layer decoder. We employed and compared different machine learning backbones, including Multilayer Perceptron (MLP), Long Short-term memory (LSTM), Gated Recurrent Unit (GRU), and Transformer. The encoder reduced the input feature dimensions to a 16-dimensional latent representation, while the decoder reconstructed the original input sequence. This reconstruction task enhanced the learning of effective temporal representations for patients, which used mean squared error (MSE) as the loss function. Furthermore, the learned latent representation was fed into an MLP for classification (Fig. 1C). Through this classification task, we can stratify the latent representations learned by the encoder based on sex during training. For this task, we utilized the binary cross-entropy (BCELoss) as the loss function and the area under the receiver operating characteristic curve (AUROC) as the metric for evaluating predictive performance. The model was trained jointly using the Adam optimizer, summing the MSE and BCELoss for optimization.

We further performed an ablation study to evaluate the impact of subsequence length and selection of phenotype categories on model performance. Specifically, we tested various time intervals (i.e., 3-, 6-, 9-, and 12-month) for constructing patient subsequence, each containing longitudinal clinical records within the specified window. To assess the influence of input features, we examined different combinations of phenotype categories: (1) demographics and vital signs, (2) demographics and diagnoses, (3) demographics and medication, and (4) demographics and lab results. Demographic variables were included in all combinations due to their largely invariant nature, whereas the other categories represent dynamic clinical variables that are more likely to influence model performance. To validate the model’s ability to capture sex-specific patterns in AD progression, we employed t-distributed stochastic neighbor embedding (t-SNE) to visualize the distribution of patients’ features at the subsequence level before and after representation learning.

### Clustering sex-specific latent representation of patients with AD

We further utilized different clustering algorithms (i.e., k-means, hierarchical agglomerative clustering, spectral clustering, and Gaussian mixture models [GMM]) on the sex-specific subsequence representations of patients with AD, aiming at identifying the clusters (i.e., disease states) of these patients with AD. To determine the optimal clustering approach for identifying distinct disease states, we evaluated the reproducibility and stability of these methods. Cluster reproducibility evaluates the extent of separation and distinctiveness among the clusters, while stability can measure whether the clusters generated by the model are stable over multiple iterations. Regarding reproducibility, we employed the silhouette score, which measures how similar a data point is to its own cluster compared to other clusters. A higher silhouette score indicates better cluster separation, where data points are closer to their own cluster center than to the neighboring clusters. Notably, if the silhouette scores are close to zero, it implies the data points are on the cluster boundaries, whereas negative scores suggest the data points are within another cluster, which is potentially incorrectly clustered. Moreover, we implemented stability testing to assess the quality of clustering by determining whether the model could consistently generate robust cluster results across multiple iterations. Specifically, we performed clustering on random subsets of the data and assessed the fluctuations in clustering outcomes using the Adjusted Rand Index (ARI), a widely used metric that quantifies the similarity between two clusters^84^. A higher ARI indicates greater consistency between clustering iterations, reflecting a more stable model. We also evaluated the stability of clustering by the Dunn index and Davies-Bouldin index. The Dunn index measures the ratio of the minimum inter-cluster distance to the maximum intra-cluster distance, with a larger value indicating better separation between clusters with higher stability. Conversely, the Davies-Bouldin index evaluates the average similarity ratio of each cluster with its most similar cluster, and a lower value suggests the clusters are more distinct and stable. To select the optimal number of clusters, the elbow method was used to identify a point where the rate of decrease in within-cluster variance slows, indicating a good trade-off between model complexity and cluster resolution.

To facilitate the observation of the distribution of identified disease states, we used principal component analysis (PCA) dimensionality reduction to visualize the distribution of the latent embeddings of patient sub-sequences. The distributions of the first two principal components were visualized using scatter plots, allowing us to qualitatively examine how sub-sequences clustered in lower-dimensional space. To quantitatively evaluate the relationship between clusters and the PCA components, we employed the Mann-Whitney 𝑈 test^85^, a non-parametric test used to compare two independent samples and assess whether they originate from the same distribution. In this context, the Mann-Whitney 𝑈 test takes the PCA component from states as parameters, compares their ranks to assess whether they come from the same distribution, and returns the test statistic and 𝑝-value to indicate statistical significance. All analyses are conducted at a significance level of 𝑝 -value < 0.05^86^, as this is a commonly used 𝑝 -value threshold for statistical significance in biomedical research. If 𝑝-value < 0.05, we reject the null hypothesis, indicating that the PCA components of sub-sequences differ significantly among states.

### Deriving sex-specific AD progression sub-phenotypes

To identify sex-specific AD progression sub-phenotypes, we first extracted the temporal representations of each subsequence from the encoder’s outputs involving sex-stratified clinical information of patients with AD. We then used the clustering algorithm to identify them into different clusters that exhibit similar temporal properties. Once the clusters of subsequences were identified, each patient’s state at a given time point was determined by the cluster assignment of the corresponding subsequence. For each patient, the identified states were concatenated chronologically along their disease progression pathways, and persistent states were merged to retain the key transitions that capture the disease progression from one state to another. For instance, we suppose that a patient has four sub-sequences containing clinical information with different time lengths (e.g., 3, 6, 9, and 12 months). Each subsequence can be grouped into a cluster (i.e., state) denoted as 𝐶_*i*_ (𝑖 = 1, 2, 3, …), assuming 3-month to 𝐶_3_, 6-month to 𝐶_4_, 9-month to 𝐶_5_, and 12-month to 𝐶_6_. The progression trajectory of this patient would be represented as “𝐶_3_ → 𝐶_4_ → 𝐶_5_ → 𝐶_6_”, which would be categorized as one of the sex-specific AD progression sub-phenotypes 𝑆_*i*_. Subsequently, all patients with AD with the same progression pathway were categorized into one sub-phenotype. To visualize the composition and state transitions of each sub-phenotype, we calculated the distribution and used the Sankey diagrams. Specifically, we quantified the number of female and male patients within each sub-phenotype and mapped the proportion of distinct disease states along their respective progression pathways. For downstream analyses, we focused only on the major sub-phenotypes that included a sufficient number of patients with AD and ensured that each state within the trajectory contributed more than 1% of the total sub-phenotype population.

### Characterization and interpretation of identified sub-phenotypes

To identify the characteristics of sex-specific AD sub-phenotypes 𝑆_*i*_, we first compared them across demographics, disease duration, prevalent comorbidities, and phenotypic features. To validate the sex specificity of the identified sub-phenotypes, we computed the Pearson correlation coefficients between sex and AD sub-phenotypes^87^. The correlation reflects the tendency for co-occurrence between each sex and sub-phenotype groups. Additionally, to facilitate the phenotypic interpretation of each sub-phenotype, we performed a detailed analysis of phenotype prevalence. For each phenotype 𝑝_*k*_, we calculated its prevalence within sub-phenotype as the proportion of patients exhibiting 𝑝_*k*_ at their latest encounter after AD onset 𝑇_*m*_. To quantify how significantly phenotypes differentiate among sub-phenotypes, we conducted pairwise chi-square tests between each sub-phenotype and all others. We then computed the average statistical significance level and the pooled odds ratio for each phenotype with respect to sub-phenotype. The average statistical significance level (the average 𝑝-value) and the pooled odds ratio were from the chi-square tests comparing phenotype 𝑝_*k*_ between 𝑆_*i*_ and other sub-phenotypes. The average statistical significance level revealed whether the prevalence of a phenotype 𝑝_*k*_ in one sub-phenotype 𝑆_*i*_ differed significantly from that in the others, while the pooled odds ratio quantified the magnitude of the difference. Moreover, to evaluate the discriminative power of the identified sex-specific AD sub-phenotypes and to identify key phenotypic predictors, we grouped female-dominant and male-dominant sub-phenotypes into pairs and employed regression models for each comparison. These models classified sub-phenotypes based on phenotypic features, and feature importance was determined by averaging the model coefficients across runs. We enhanced clinical interpretability by mapping all phenotypes to Phecode Chapters^88^, which classify phenotypes into 18 organ system categories. For visualization and downstream analyses, we further restricted our focus to the phenotypes with the highest prevalence and statistically significant differences across all sub-phenotypes (i.e., 𝑝-value < 0.05).

To gain a deeper understanding of disease progression dynamics across sex-specific AD sub-phenotypes, we performed survival analysis using the Kaplan–Meier method to estimate the time to AD onset among patients with prevalent comorbid phenotypes^89^. The objective of survival analysis was to examine whether the presence of specific comorbidities within each sub-phenotype was associated with the differences in risk and time to AD onset. To investigate this, we generated Kaplan–Meier survival curves to compare survival probabilities across sex-based AD sub-phenotypes for each comorbid condition. To statistically compare survival probability differences between sub-phenotypes, we applied the two-sided log-rank test, providing a robust evaluation of group-level differences in AD onset^90^. We further employed the Cox proportional hazards model to estimate the relative risk of AD onset for each sub-phenotype using the most prevalent group as the reference. Hazard ratios (HRs) and their corresponding 95% confidence intervals (CIs) were calculated using robust standard errors^91^ to ensure reliable inference. Additionally, we calculated the cumulative prevalence of the top three most common phenotypes within each time interval to capture comorbidity accumulation patterns. Prior studies^92–94^ have shown that rising prevalence of certain comorbidities may be associated with the onset of AD. Therefore, we compared the trajectory of AD prevalence of AD against the accumulation of comorbid conditions across different sub-phenotypes. While Kaplan–Meier curves can depict the decline in survival probability from the index date to AD diagnosis, cumulative prevalence curves reveal the temporal patterns of comorbidity accumulation relative to AD onset across sub-phenotypes. Together, these analyses provide valuable insights into the temporal dynamics and risk profiles of sex-specific AD progression sub-phenotypes, identifying potential time windows for early intervention based on comorbidity patterns.

## Data availability

The OneFlorida+ Data Trust is accessible to investigators for research purposes through formal policies and procedures established by OneFlorida+ Executive Committee. Researchers can initiate and complete Prep-to-Research Data Query via the OneFlorida+ Application Portal (https://webportalapp.com/sp/login/onefl_preptoresearch). OneFlorida+ Informatics for Integrating Biology and the Bedside (i2b2) (https://onefl.net/front-door/onefli2b2/) facilitates cohort discovery and conducts descriptive analyses, which provide aggregate patient counts, sociodemographic and health characteristics, and healthcare practice demographics. Access to individual-level data requires a formal data request submission. For information on data access procedures, investigators can schedule a consultation (https://onefl.net/front-door/consultation/).

## Code availability

The codes for this study are publicly available at https://github.com/UF-HOBI-Yin-Lab/AD_sex_subtype.

## ACKNOWLEDGMENTS

This study was partially supported by grants from the Centers for Disease Control and Prevention (1U18DP006512), the National Institute of Environmental Health Sciences (R21ES032762), and the NIH National Center for Advancing Translational Sciences (UL1TR001427).

## Contributions

R.Y. conceived ideas. W.M., Q.Y., and R.Y. designed the experiments. W.M. and Q.Y. performed the experiments. R.Y. and J.X. developed the initial concept. W.M. wrote the initial manuscript with support from R.Y. Q.Y., J.X., Y.H., Q.S., C.W., A.M., Q.M., L.S., J.B., and R.Y. provided critical feedback and helped shape the research, analysis, and manuscript. R.Y. supervised the project.

## Ethics declarations

Competing interests

The authors declare no competing interests.

## Supplementary information

<u>Supplementary Information</u>

<u>Description of Additional Supplementary Files</u>

<u>Supplementary Data 1</u>

<u>Supplementary Data 2</u>

## Notes

### Competing Interest Statement

The authors have declared no competing interest.

### Funding Statement

This study was partially supported by grants from Centers for Disease Control and Prevention (1U18DP006512), National Institute of Environmental Health Sciences (R21ES032762) and the NIH National Center for Advancing Translational Sciences (UL1TR001427).

### Author Declarations

Ethics committee/IRB of University of Florida gave ethical approval for this work.

### Summary of Updates

1. Title changed. 2. Added an author Dr. Qiang Yang. 3. Abstract modified. 4. Added Experiments and changed all figures.

## References

1. 2024 Alzheimer’s disease facts and figures. Alzheimers. Dement. 20, 3708–3821 (2024).

2. Rajan, K. B. et al. Population estimate of people with clinical Alzheimer’s disease and mild cognitive impairment in the United States (2020-2060). Alzheimers. Dement. 17, 1966–1975 (2021).

3. Sperling, R. A. et al. Toward defining the preclinical stages of Alzheimer’s disease: recommendations from the National Institute on Aging-Alzheimer’s Association workgroups on diagnostic guidelines for Alzheimer’s disease. Alzheimers. Dement. 7, 280–292 (2011).

4. Albert, M. S. et al. The diagnosis of mild cognitive impairment due to Alzheimer’s disease: Recommendations from the national institute on aging-Alzheimer’s association workgroups on diagnostic guidelines for Alzheimer’s disease. Focus (Am. Psychiatr. Publ.) 11, 96–106 (2013).

5. Sun, B.-L. et al. Clinical research on Alzheimer’s disease: Progress and perspectives. Neurosci. Bull. 34, 1111–1118 (2018).

6. Kim, Y., Lhatoo, S., Zhang, G.-Q., Chen, L. & Jiang, X. Temporal phenotyping for transitional disease progress: An application to epilepsy and Alzheimer’s disease. J. Biomed. Inform. 107, 103462 (2020).

7. Li, J.-Q. et al. Risk factors for predicting progression from mild cognitive impairment to Alzheimer’s disease: a systematic review and meta-analysis of cohort studies. J. Neurol. Neurosurg. Psychiatry 87, 476–484 (2016).

8. Salvadó, G. et al. Disease staging of Alzheimer’s disease using a CSF-based biomarker model. *Nat*. Aging 4, 694–708 (2024).

9. Jamalian, S. et al. Modeling Alzheimer’s disease progression utilizing clinical trial and ADNI data to predict longitudinal trajectory of CDR-SB. CPT Pharmacometrics Syst. Pharmacol. 12, 1029–1042 (2023).

10. Stallard, E. et al. Estimation and validation of a multiattribute model of Alzheimer disease progression. Med. Decis. Making 30, 625–638 (2010).

11. Lam, B., Masellis, M., Freedman, M., Stuss, D. T. & Black, S. E. Clinical, imaging, and pathological heterogeneity of the Alzheimer’s disease syndrome. Alzheimers. Res. Ther. 5, 1 (2013).

12. Goyal, D. et al. Characterizing heterogeneity in the progression of Alzheimer’s disease using longitudinal clinical and neuroimaging biomarkers. Alzheimers Dement. (Amst.) 10, 629–637 (2018).

13. Young-Pearse, T. L., Lee, H., Hsieh, Y.-C., Chou, V. & Selkoe, D. J. Moving beyond amyloid and tau to capture the biological heterogeneity of Alzheimer’s disease. Trends Neurosci. 46, 426–444 (2023).

14. Dumitrescu, L., Mayeda, E. R., Sharman, K., Moore, A. M. & Hohman, T. J. Sex Differences in the Genetic Architecture of Alzheimer’s Disease. Curr. Genet. Med. Rep. 7, 13–21 (2019).

15. Dumitrescu, L. et al. Sex differences in the genetic predictors of Alzheimer’s pathology. Brain 142, 2581–2589 (2019).

16. Zhu, D., Montagne, A. & Zhao, Z. Alzheimer’s pathogenic mechanisms and underlying sex difference. Cell. Mol. Life Sci. 78, 4907–4920 (2021).

17. Guo, L., Zhong, M. B., Zhang, L., Zhang, B. & Cai, D. Sex Differences in Alzheimer’s Disease: Insights From the Multiomics Landscape. Biol. Psychiatry 91, 61–71 (2022).

18. Barnes, L. L. et al. Sex differences in the clinical manifestations of Alzheimer disease pathology. Arch. Gen. Psychiatry 62, 685–691 (2005).

19. Fan, C. C. et al. Sex-dependent autosomal effects on clinical progression of Alzheimer’s disease. Brain 143, 2272–2280 (2020).

20. Kamizato, C., Osawa, A., Maeshima, S., Kagaya, H. & Arai, H. Activity level by clinical severity and sex differences in patients with Alzheimer disease and mild cognitive impairment. Psychogeriatrics 23, 815–820 (2023).

21. Oveisgharan, S. et al. Sex differences in Alzheimer’s disease and common neuropathologies of aging. Acta Neuropathol. 136, 887–900 (2018).

22. Ullah, M. F. et al. Impact of sex differences and gender specificity on behavioral characteristics and pathophysiology of neurodegenerative disorders. Neurosci. Biobehav. Rev. 102, 95–105 (2019).

23. Eissman, J. M. et al. Sex differences in the genetic architecture of cognitive resilience to Alzheimer’s disease. Brain 145, 2541–2554 (2022).

24. Chêne, G. et al. Gender and incidence of dementia in the Framingham Heart Study from mid-adult life. Alzheimers. Dement. 11, 310–320 (2015).

25. Farrer, L. A. Effects of age, sex, and ethnicity on the association between apolipoprotein E genotype and Alzheimer disease. JAMA 278, 1349 (1997).

26. Altmann, A., Tian, L., Henderson, V. W., Greicius, M. D. & Alzheimer’s Disease Neuroimaging Initiative Investigators. Sex modifies the APOE-related risk of developing Alzheimer disease. Ann. Neurol. 75, 563–573 (2014).

27. Kim, J., Basak, J. M. & Holtzman, D. M. The role of apolipoprotein E in Alzheimer’s disease. Neuron 63, 287–303 (2009).

28. Ungar, L., Altmann, A. & Greicius, M. D. Apolipoprotein E, gender, and Alzheimer’s disease: an overlooked, but potent and promising interaction. Brain Imaging Behav. 8, 262–273 (2014).

29. Mosconi, L. et al. Sex differences in Alzheimer risk: Brain imaging of endocrine vs chronologic aging. Neurology 89, 1382–1390 (2017).

30. Belloy, M. E. et al. APOE genotype and Alzheimer disease risk across age, sex, and population ancestry. JAMA neurology vol. 80 1284–1294 (2023).

31. Hohman, T. J. et al. Sex-specific association of apolipoprotein E with cerebrospinal fluid levels of tau. JAMA Neurol. 75, 989 (2018).

32. Goldstein, J. M., Langer, A. & Lesser, J. A. Sex differences in disorders of the brain and heart—A global crisis of multimorbidity and novel opportunity. JAMA Psychiatry 78, 7 (2021).

33. Ossenkoppele, R. et al. Assessment of demographic, genetic, and imaging variables associated with brain resilience and cognitive resilience to pathological tau in patients with Alzheimer disease. JAMA Neurol. 77, 632–642 (2020).

34. Digma, L. A. et al. Women can bear a bigger burden: ante- and post-mortem evidence for reserve in the face of tau. Brain Commun. 2, fcaa025 (2020).

35. Thalhauser, C. J. & Komarova, N. L. Alzheimer’s disease: rapid and slow progression. J. R. Soc. Interface 9, 119–126 (2012).

36. Alexander, N., Alexander, D. C., Barkhof, F. & Denaxas, S. Identifying and evaluating clinical subtypes of Alzheimer’s disease in care electronic health records using unsupervised machine learning. BMC Med. Inform. Decis. Mak. 21, 343 (2021).

37. van der Haar, D., Moustafa, A., Warren, S. L., Alashwal, H. & van Zyl, T. An Alzheimer’s disease category progression sub-grouping analysis using manifold learning on ADNI. Sci. Rep. 13, 10483 (2023).

38. Murray, M. E. et al. Neuropathologically defined subtypes of Alzheimer’s disease with distinct clinical characteristics: a retrospective study. Lancet Neurol. 10, 785–796 (2011).

39. Barbieri, C., Neri, L., Stuard, S., Mari, F. & Martín-Guerrero, J. D. From electronic health records to clinical management systems: how the digital transformation can support healthcare services. Clin. Kidney J. 16, 1878–1884 (2023).

40. Woldemariam, M. T. & Jimma, W. Adoption of electronic health record systems to enhance the quality of healthcare in low-income countries: a systematic review. BMJ Health Care Inform. 30, e100704 (2023).

41. Meng, W. et al. An interpretable population graph network to identify rapid progression of Alzheimer’s disease using UK Biobank. medRxiv (2024) doi:10.1101/2024.03.27.24304966.

42. Xu, J. et al. Identification of Outcome-Oriented Progression Subtypes from Mild Cognitive Impairment to Alzheimer’s Disease Using Electronic Health Records. medRxiv (2023) doi:10.1101/2023.07.27.23293270.

43. Wu, P.-F. et al. Growth differentiation factor 15 is associated with Alzheimer’s disease risk. Front. Genet. 12, 700371 (2021).

44. Lee, C. & van der Schaar, M. Temporal phenotyping using deep predictive clustering of disease progression. arXiv [physics.med-ph] (2020).

45. Hochreiter, S. & Schmidhuber, J. Long short-term memory. Neural Comput. 9, 1735–1780 (1997).

46. Tang, A. S. et al. Deep phenotyping of Alzheimer’s disease leveraging electronic medical records identifies sex-specific clinical associations. Nat. Commun. 13, 675 (2022).

47. Tang, A. S. et al. Leveraging electronic health records and knowledge networks for Alzheimer’s disease prediction and sex-specific biological insights. *Nat*. Aging 4, 379–395 (2024).

48. Vaswani, A., et al. Attention is all you need. *arXiv [cs.CL]* (2017).

49. Chung, J., Gulcehre, C., Cho, K. & Bengio, Y. Empirical evaluation of gated recurrent neural networks on sequence modeling. arXiv [cs.NE] (2014).

50. Bengio, Y., Schwenk, H., Senécal, J.-S., Morin, F. & Gauvain, J.-L. Neural probabilistic language models. in Innovations in Machine Learning 137–186 (Springer-Verlag, Berlin/Heidelberg, 2006).

51. van der Maaten, L. & Hinton, G. Visualizing Data using t-SNE. J. Mach. Learn. Res. 9, 2579–2605 (2008).

52. Hartigan, J. A. & Wong, M. A. Journal of the royal statistical society. series c (applied statistics). 28, 100–108 (1979).

53. Murtagh, F. & Legendre, P. Ward’s hierarchical agglomerative clustering method: Which algorithms implement ward’s criterion? J. Classif. 31, 274–295 (2014).

54. Zare, H., Shooshtari, P., Gupta, A. & Brinkman, R. R. Data reduction for spectral clustering to analyze high throughput flow cytometry data. BMC Bioinformatics 11, 403 (2010).

55. Reynolds, D. Gaussian mixture models. in Encyclopedia of Biometrics 659–663 (Springer US, Boston, MA, 2009).

56. Dunn, J. C. A fuzzy relative of the ISODATA process and its use in detecting compact well-separated clusters. J. Cybern. 3, 32–57 (1973).

57. Pearson, K. X. On the criterion that a given system of deviations from the probable in the case of a correlated system of variables is such that it can be reasonably supposed to have arisen from random sampling. Lond. Edinb. Dublin Philos. Mag. J. Sci. 50, 157–175 (1900).

58. Kaplan, E. L. & Meier, P. Nonparametric Estimation from Incomplete Observations. J. Am. Stat. Assoc. 53, 457 (1958).

59. Cox, D. R. Regression models and life-tables. J. R. Stat. Soc. Series B Stat. Methodol. 34, 187–202 (1972).

60. With Chartbook on Long-Term Trends in Health. (United States; Hyattsville, MD).

61. Iturria-Medina, Y. et al. Early role of vascular dysregulation on late-onset Alzheimer’s disease based on multifactorial data-driven analysis. Nat. Commun. 7, 11934 (2016).

62. Nucera, A. & Hachinski, V. Cerebrovascular and Alzheimer disease: fellow travelers or partners in crime? J. Neurochem. 144, 513–516 (2018).

63. de la Torre, J. C. Alzheimer disease as a vascular disorder. Stroke 33, 1152–1162 (2002).

64. Attems, J. & Jellinger, K. A. The overlap between vascular disease and Alzheimer’s disease - lessons from pathology. BMC Med. 12, (2014).

65. Heppner, F. L., Ransohoff, R. M. & Becher, B. Immune attack: the role of inflammation in Alzheimer disease. Nat. Rev. Neurosci. 16, 358–372 (2015).

66. Khosroazad, S. et al. Sleep movements and respiratory coupling as a biobehavioral metric for early Alzheimer’s disease in independently dwelling adults. BMC Geriatr. 23, 252 (2023).

67. Jankowska-Kieltyka, M., Roman, A. & Nalepa, I. The air we breathe: Air pollution as a prevalent proinflammatory stimulus contributing to neurodegeneration. Front. Cell. Neurosci. 15, 647643 (2021).

68. Honjo, K., van Reekum, R. & Verhoeff, N. P. L. G. Alzheimer’s disease and infection: do infectious agents contribute to progression of Alzheimer’s disease? Alzheimers. Dement. 5, 348–360 (2009).

69. Goveas, J. S., Espeland, M. A., Woods, N. F., Wassertheil-Smoller, S. & Kotchen, J. M. Depressive symptoms and incidence of mild cognitive impairment and probable dementia in elderly women: The Women’s Health Initiative Memory Study: depression and incident MCI and dementia. J. Am. Geriatr. Soc 59, 57–66 (2011).

70. Sanacora, G., Zarate, C. A., Krystal, J. H. & Manji, H. K. Targeting the glutamatergic system to develop novel, improved therapeutics for mood disorders. Nat. Rev. Drug Discov. 7, 426–437 (2008).

71. Eriksson, I., Gustafson, Y., Fagerström, L. & Olofsson, B. Urinary tract infection in very old women is associated with delirium. Int. Psychogeriatr. 23, 496–502 (2011).

72. Yuan, S. et al. Digestive system diseases, genetic risk, and incident dementia: A prospective cohort study. Am. J. Prev. Med. 66, 516–525 (2024).

73. Jang, Y. J. et al. Additive interaction of mid-to late-life depression and cerebrovascular disease on the risk of dementia: a nationwide population-based cohort study. Alzheimer’s research & therapy 13, 1– 3 (2021).

74. Xu, J. et al. Federated Learning for Healthcare Informatics. Int. J. Healthc. Inf. Syst. Inform. 5, 1–19 (2021).

75. Integrative Analyses of Multimodal Clinical Neuroimaging,Genetic,and Data Identify Subtypes and Potential Treatments for Heterogeneous Parkinson Disease.

76. Shenkman, E. et al. OneFlorida Clinical Research Consortium: Linking a Clinical and Translational Science Institute With a Community-Based Distributive Medical Education Model. Acad. Med. 93, 451–455 (2018).

77. Hogan, W. R. et al. The OneFlorida Data Trust: a centralized, translational research data infrastructure of statewide scope. J. Am. Med. Inform. Assoc. 29, 686–693 (2022).

78. OneFlorida+ clinical research network. https://onefl.net/.

79. Denny, J. C. et al. PheWAS: demonstrating the feasibility of a phenome-wide scan to discover gene-disease associations. Bioinformatics 26, 1205–1210 (2010).

80. Dubois, B., von Arnim, C. A. F., Burnie, N., Bozeat, S. & Cummings, J. Biomarkers in Alzheimer’s disease: role in early and differential diagnosis and recognition of atypical variants. Alzheimers. Res. Ther. 15, 175 (2023).

81. WHOCC. ATCDDD - Home. https://atcddd.fhi.no/.

82. LOINC to CPT mapping. (2006).

83. Van Buuren, S. & Groothuis-Oudshoorn, K. Multivariate imputation by chained equations in R. Journal of statistical software 45, 1–67 (2011).

84. Rand, W. M. Objective criteria for the evaluation of clustering methods. J. Am. Stat. Assoc. 66, 846 (1971).

85. Mann, H. B. & Whitney, D. R. On a Test of Whether one of Two Random Variables is Stochastically Larger than the Other. Ann. Math. Stat. 18, 50–60 (1947).

86. Gale, R. P., Hochhaus, A. & Zhang, M.-J. What is the (p-) value of the P-value? Leukemia 30, 1965– 1967 (2016).

87. Schober, P., Boer, C. & Schwarte, L. A. Correlation coefficients: Appropriate use and interpretation. Anesth. Analg. 126, 1763–1768 (2018).

88. Bastarache, L. Using phecodes for research with the electronic health record: From PheWAS to PheRS. Annu. Rev. Biomed. Data Sci. 4, 1–19 (2021).

89. Elton, R. A. & Lee, E. T. Statistical methods for survival data analysis. Biometrics 51, 383 (1995).

90. Mantel, N. Evaluation of survival data and two new rank order statistics arising in its consideration.Cancer Chemother. Rep. 50, 163–170 (1966).

91. Breslow, N. E. Analysis of survival data under the proportional hazards model. Int. Stat. Rev. 43, 45 (1975).

92. Spina, S. et al. Comorbid neuropathological diagnoses in early versus late-onset Alzheimer’s disease. Brain 144, 2186–2198 (2021).

93. Santiago, J. A. & Potashkin, J. A. The impact of disease comorbidities in Alzheimer’s disease. Front. Aging Neurosci. 13, 631770 (2021).

94. Butler, L. M., Houghton, R., Abraham, A., Vassilaki, M. & Durán-Pacheco, G. Comorbidity trajectories associated with Alzheimer’s disease: A matched case-control study in a United States claims database. Front. Neurosci. 15, 749305 (2021).

